# Largest GWAS (N=1,126,563) of Alzheimer’s Disease Implicates Microglia and Immune Cells

**DOI:** 10.1101/2020.11.20.20235275

**Authors:** Douglas P Wightman, Iris E Jansen, Jeanne E. Savage, Alexey A Shadrin, Shahram Bahrami, Arvid Rongve, Sigrid Børte, Bendik S Winsvold, Ole Kristian Drange, Amy E Martinsen, Anne Heidi Skogholt, Cristen Willer, Geir Bråthen, Ingunn Bosnes, Jonas Bille Nielsen, Lars Fritsche, Laurent F. Thomas, Linda M Pedersen, Maiken E Gabrielsen, Marianne Bakke Johnsen, Tore Wergeland Meisingset, Wei Zhou, Petra Proitsi, Angela Hodges, Richard Dobson, Latha Velayudhan, 23andMe Research Team, Julia M Sealock, Lea K Davis, Nancy L. Pedersen, Chandra A. Reynolds, Ida K. Karlsson, Sigurdur Magnusson, Hreinn Stefansson, Steinunn Thordardottir, Palmi V. Jonsson, Jon Snaedal, Anna Zettergren, Ingmar Skoog, Silke Kern, Margda Waern, Henrik Zetterberg, Kaj Blennow, Eystein Stordal, Kristian Hveem, John-Anker Zwart, Lavinia Athanasiu, Ingvild Saltvedt, Sigrid B Sando, Ingun Ulstein, Srdjan Djurovic, Tormod Fladby, Dag Aarsland, Geir Selbæk, Stephan Ripke, Kari Stefansson, Ole A. Andreassen, Danielle Posthuma

## Abstract

Late-onset Alzheimer’s disease is a prevalent age-related polygenic disease that accounts for 50-70% of dementia cases^1^. Late-onset Alzheimer’s disease is caused by a combination of many genetic variants with small effect sizes and environmental influences. Currently, only a fraction of the genetic variants underlying Alzheimer’s disease have been identified^2,3^. Here we show that increased sample sizes allowed for identification of seven novel genetic loci contributing to Alzheimer’s disease. We highlighted eight potentially causal genes where gene expression changes are likely to explain the association. Human microglia were found as the only cell type where the gene expression pattern was significantly associated with the Alzheimer’s disease association signal. Gene set analysis identified four independent pathways for associated variants to influence disease pathology. Our results support the importance of microglia, amyloid and tau aggregation, and immune response in Alzheimer’s disease. We anticipate that through collaboration the results from this study can be included in larger meta-analyses of Alzheimer’s disease to identify further genetic variants which contribute to Alzheimer’s pathology. Furthermore, the increased understanding of the mechanisms that mediate the effect of genetic variants on disease progression will help identify potential pathways and gene-sets as targets for drug development.

## Main text

Dementia has an age- and sex-standardised prevalence of ∼7.1% in Europeans^4^, with Alzheimer’s disease (AD) being the most common form of dementia (50-70% of cases)^1^. AD is pathologically characterized by the presence of amyloid-beta plaques and tau neurofibrillary tangles in the brain^5^. Most patients are diagnosed with AD after the age of 65, termed late onset AD (LOAD), while only 1% of the AD cases have an early onset (before the age of 65)^5^. Based on twin studies, the heritability of LOAD is estimated to be ∼60-80%^6,7^, suggesting that a large proportion of individual differences in LOAD risk is driven by genetics. The heritability of LOAD is spread across many genetic variants; however, Zhang *et al*. (2020)^2^ suggested that LOAD is more of an oligogenic than polygenic disorder due to the large effects of *APOE* variants. According to Zhang *et al*. (2020) and Holland *et al*. (2020)^3^ there are predicted to be ∼100-1000 causal variants contributing to LOAD and only a fraction have been identified. Increasing the sample size of GWAS studies will improve the statistical power to identify the missing causal variants and may highlight novel disease mechanisms.

The largest previous GWAS of LOAD, performed in 2019, identified 29 risk loci from 71,880 (46,613 proxy) cases and 383,378 (318,246 proxy) controls^8^. Our current study expands this to include 90,338 (46,613 proxy) cases and 1,036,225 (318,246 proxy) controls. The recruitment of LOAD cases can be difficult due to the late age of onset so proxy cases can allow for the inclusion of younger individuals by estimating their risk of LOAD using parental status. In the current study, we identified 38 loci, including seven loci that have not been reported previously. Extensive functional follow-up analyses implicated tissues, cell types, and genes of interest through tissue and cell type enrichment, colocalization, and statistical fine-mapping. This study highlights microglia, immune cells, and protein catabolism as relevant to LOAD while identifying novel genes of potential interest.

### Genome-wide inferences

We meta-analyzed data from 13 cohorts, totaling 1,126,563 individuals (**Supplementary Table 1**). Proxy cases and controls were defined based on known parental LOAD status weighted by parental age (**Online Methods**). The meta-analysis identified 3915 significant (*P*< 5×10^−8^) variants across 38 independent loci (**Table 1, Figure 1**). Of those 38 loci, seven have not shown associations with LOAD in previous GWAS, and five of those novel loci have not been associated with any form of dementia. Three of the seven novel loci are inconclusive due to low support from surrounding variants (**Supplementary Results**). In comparison to our previous meta-analysis^8^, we failed to replicate the association of *ADAMTS4, HESX1, CNTNAP2, KAT8, SCIMP, ALPK2*, and *AC074212*.*3* in the current study.

**Table 1:**
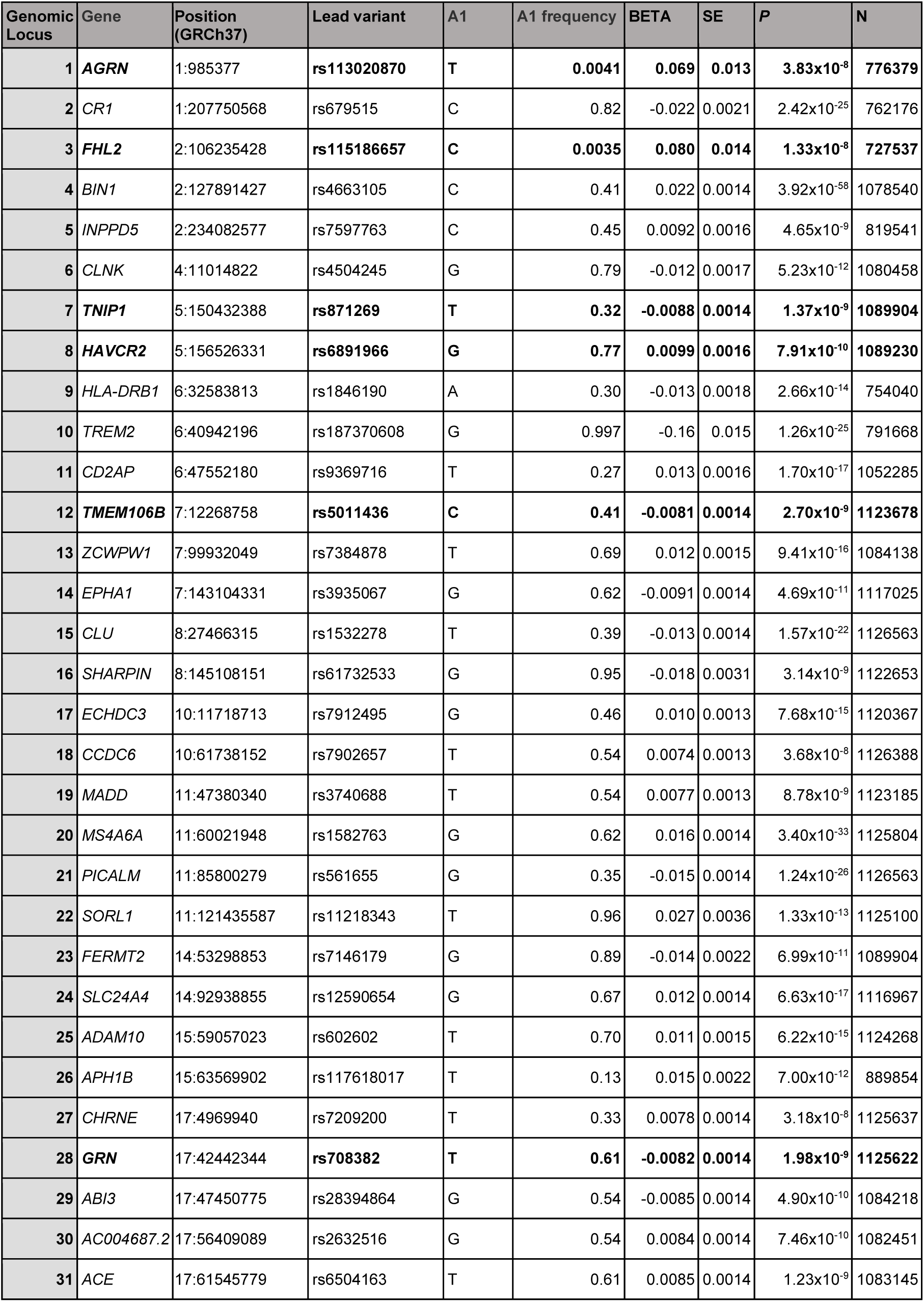
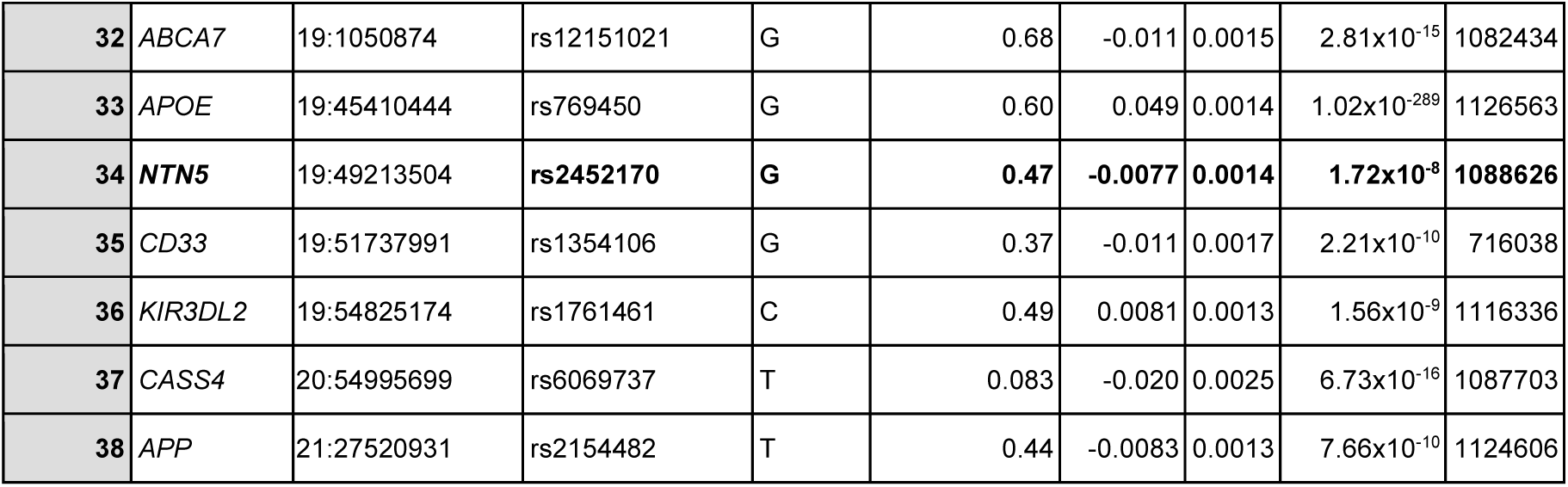
The 38 genomic risk loci identified from 90,338 (46,613 proxy) cases and 1,036,225 (318,246 proxy) controls. The novel loci are highlighted in bold. The genes were assigned based on colocalization results, fine-mapping results, and previous literature.

| Genomic Locus | Gene | Position (GRCh37) | Lead variant | A1 | A1 frequency | BETA | SE | P | N |
| --- | --- | --- | --- | --- | --- | --- | --- | --- | --- |
| 1 | **AGRN** | 1:985377 | **rs113020870** | T | 0.0041 | 0.069 | 0.013 | 3.83x10<sup>-8</sup> | 776379 |
| 2 | CR1 | 1:207750568 | rs679515 | C | 0.82 | -0.022 | 0.0021 | 2.42x10<sup>-25</sup> | 762176 |
| 3 | **FHL2** | 2:106235428 | **rs115186657** | C | 0.0035 | 0.080 | 0.014 | 1.33x10<sup>-8</sup> | 727537 |
| 4 | BIN1 | 2:127891427 | rs4663105 | C | 0.41 | 0.022 | 0.0014 | 3.92x10<sup>-58</sup> | 1078540 |
| 5 | INPPD5 | 2:234082577 | rs7597763 | C | 0.45 | 0.0092 | 0.0016 | 4.65x10<sup>-9</sup> | 819541 |
| 6 | CLNK | 4:11014822 | rs4504245 | G | 0.79 | -0.012 | 0.0017 | 5.23x10<sup>-12</sup> | 1080458 |
| 7 | **TNIP1** | 5:150432388 | **rs871269** | T | 0.32 | -0.0088 | 0.0014 | 1.37x10<sup>-9</sup> | 1089904 |
| 8 | **HAVCR2** | 5:156526331 | **rs6891966** | G | 0.77 | 0.0099 | 0.0016 | 7.91x10<sup>-10</sup> | 1089230 |
| 9 | HLA-DRB1 | 6:32583813 | rs1846190 | A | 0.30 | -0.013 | 0.0018 | 2.66x10<sup>-14</sup> | 754040 |
| 10 | TREM2 | 6:40942196 | rs187370608 | G | 0.997 | -0.16 | 0.015 | 1.26x10<sup>-25</sup> | 791668 |
| 11 | CD2AP | 6:47552180 | rs9369716 | T | 0.27 | 0.013 | 0.0016 | 1.70x10<sup>-17</sup> | 1052285 |
| 12 | **TMEM106B** | 7:12268758 | **rs5011436** | C | 0.41 | -0.0081 | 0.0014 | 2.70x10<sup>-9</sup> | 1123678 |
| 13 | ZCWPW1 | 7:99932049 | rs7384878 | T | 0.69 | 0.012 | 0.0015 | 9.41x10<sup>-16</sup> | 1084138 |
| 14 | EPHA1 | 7:143104331 | rs3935067 | G | 0.62 | -0.0091 | 0.0014 | 4.69x10<sup>-11</sup> | 1117025 |
| 15 | CLU | 8:27466315 | rs1532278 | T | 0.39 | -0.013 | 0.0014 | 1.57x10<sup>-22</sup> | 1126563 |
| 16 | SHARPIN | 8:145108151 | rs61732533 | G | 0.95 | -0.018 | 0.0031 | 3.14x10<sup>-9</sup> | 1122653 |
| 17 | ECHDC3 | 10:11718713 | rs7912495 | G | 0.46 | 0.010 | 0.0013 | 7.68x10<sup>-15</sup> | 1120367 |
| 18 | CCDC6 | 10:61738152 | rs7902657 | T | 0.54 | 0.0074 | 0.0013 | 3.68x10<sup>-8</sup> | 1126388 |
| 19 | MADD | 11:47380340 | rs3740688 | T | 0.54 | 0.0077 | 0.0013 | 8.78x10<sup>-9</sup> | 1123185 |
| 20 | MS4A6A | 11:60021948 | rs1582763 | G | 0.62 | 0.016 | 0.0014 | 3.40x10<sup>-33</sup> | 1125804 |
| 21 | PICALM | 11:85800279 | rs561655 | G | 0.35 | -0.015 | 0.0014 | 1.24x10<sup>-26</sup> | 1126563 |
| 22 | SORL1 | 11:121435587 | rs11218343 | T | 0.96 | 0.027 | 0.0036 | 1.33x10<sup>-13</sup> | 1125100 |
| 23 | FERMT2 | 14:53298853 | rs7146179 | G | 0.89 | -0.014 | 0.0022 | 6.99x10<sup>-11</sup> | 1089904 |
| 24 | SLC24A4 | 14:92938855 | rs12590654 | G | 0.67 | 0.012 | 0.0014 | 6.63x10<sup>-17</sup> | 1116967 |
| 25 | ADAM10 | 15:59057023 | rs602602 | T | 0.70 | 0.011 | 0.0015 | 6.22x10<sup>-15</sup> | 1124268 |
| 26 | APH1B | 15:63569902 | rs117618017 | T | 0.13 | 0.015 | 0.0022 | 7.00x10<sup>-12</sup> | 889854 |
| 27 | CHRNE | 17:4969940 | rs7209200 | T | 0.33 | 0.0078 | 0.0014 | 3.18x10<sup>-8</sup> | 1125637 |
| 28 | **GRN** | 17:42442344 | **rs708382** | T | 0.61 | -0.0082 | 0.0014 | 1.98x10<sup>-9</sup> | 1125622 |
| 29 | ABI3 | 17:47450775 | rs28394864 | G | 0.54 | -0.0085 | 0.0014 | 4.90x10<sup>-10</sup> | 1084218 |
| 30 | AC004687.2 | 17:56409089 | rs2632516 | G | 0.54 | 0.0084 | 0.0014 | 7.46x10<sup>-10</sup> | 1082451 |
| 31 | ACE | 17:61545779 | rs6504163 | T | 0.61 | 0.0085 | 0.0014 | 1.23x10<sup>-9</sup> | 1083145 |
| 32 | ABCA7 | 19:1050874 | rs12151021 | G | 0.68 | -0.011 | 0.0015 | 2.81x10<sup>-15</sup> | 1082434 |
| 33 | APOE | 19:45410444 | rs769450 | G | 0.60 | 0.049 | 0.0014 | 1.02x10<sup>-289</sup> | 1126563 |
| 34 | **NTN5** | 19:49213504 | **rs2452170** | **G** | **0.47** | **-0.0077** | **0.0014** | **1.72x10<sup>-8</sup>** | **1088626** |
| 35 | CD33 | 19:51737991 | rs1354106 | G | 0.37 | -0.011 | 0.0017 | 2.21x10<sup>-10</sup> | 716038 |
| 36 | KIR3DL2 | 19:54825174 | rs1761461 | C | 0.49 | 0.0081 | 0.0013 | 1.56x10<sup>-9</sup> | 1116336 |
| 37 | CASS4 | 20:54995699 | rs6069737 | T | 0.083 | -0.020 | 0.0025 | 6.73x10<sup>-16</sup> | 1087703 |
| 38 | APP | 21:27520931 | rs2154482 | T | 0.44 | -0.0083 | 0.0013 | 7.66x10<sup>-10</sup> | 1124606 |

**Figure 1:**
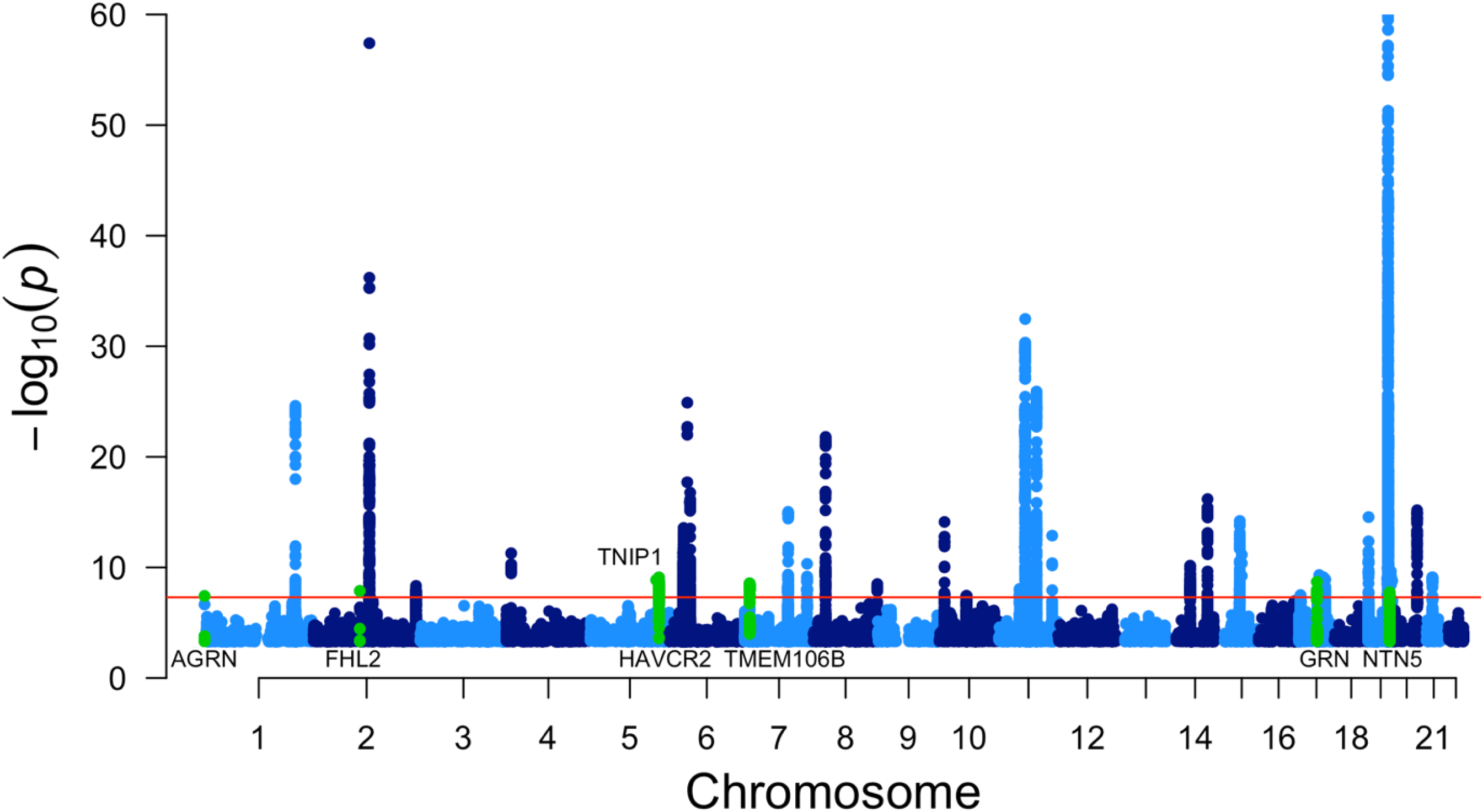
A Manhattan plot of the meta-analysis results highlighting 38 loci, including 7 novel regions. Only variants with a *P*< 0.0005 are displayed. The *APOE* region cannot be fully observed because the y-axis is limited to the top variant in the second most significant locus, *-log10*(1×10^−60^), in order to display the less significant variants. The red line represents genome wide significance (5×10^−8^). The novel loci are highlighted in green and indicated by the assigned gene name.

One novel locus (*NTN5* locus) was relatively close to the *APOE* locus (within 2.7Mb) and thus could be influenced by the strong association signal of *APOE*. However, we used GCTA-COJO^9^ to identify independently associated variants and found the *NTN5* region to be unaffected when conditioning on the *APOE* region (**Supplementary Results**), suggesting this *NTN5* locus is a LOAD risk factor independent from *APOE*.

The liability-scale SNP heritability was estimated by linkage disequilibrium score (LDSC) regression^10^ to be 0.025 (SE=0.0043) given a population prevalence of 0.05 (*APOE* region excluded). This estimate is low but similar to the estimate obtained in a previous GWAS meta-analysis (h_2l_=0.055,SE=0.0099)^8^. The genetic correlation^11^ between proxy LOAD and case-control LOAD was estimated at 0.83 (SE=0.21, *P*=6.61×10^−5^). Separate Manhattan plots for the LOAD proxy data and the case-control LOAD data are available in the Supplementary Results (**Supplementary Figures 1 & 2**). Across 855 external phenotypes in LDhub^12^, two significant genetic correlations with the meta-analysis results were observed (**Supplementary Table 2**). The strongest correlation was with a previous LOAD study conducted by Lambert *et al*. (2013)^13^ (r_g_=1.18, SE=0.19, *P*_*Bonferroni*_=2.42×10^−7^). The other significant correlation was with the UK Biobank (UKB)^14^ trait “Illnesses of mother: Alzheimer’s disease/dementia” (r_g_=0.80, SE=0.11, *P*_*Bonferroni*_=6.38×10^−10^). The current study included individuals which are also included in Lambert *et al*. (2013)^13^ and the UKB.

### Tissue type, cell type, and gene set enrichment

MAGMA tissue specificity analysis^15^ identified spleen (*P*_*bonferroni*_=0.034) as the GTEx tissue where expression of the significant MAGMA genes was enriched (**Supplementary Figure 2, Supplementary Table 3**). Spleen was also significant in the previous MAGMA tissue specificity analysis performed in Jansen *et al*. (2019)^8^ and is a known contributor to immune function. To investigate enrichment at the cell type level, FUMA cell type analysis^16^ was performed with a collection of cell types in mouse brain, human brain, and human blood tissue, resulting in 6 single-cell (scRNA-seq) datasets significantly associated, after multiple testing correction (*P*<5.39×10^−5^), with the expression of LOAD-associated genes (**Supplementary Figure 4, Supplementary Table 4**). The only significant cell type in all six independent scRNA datasets was microglia. Human microglia have not been previously identified in cell type analysis for LOAD. A combination of the cell type and tissue specificity results identifies microglia and immune tissues as potential experimental models for identifying the contribution of LOAD-associated genes towards LOAD pathogenesis.

**Figure 2:**
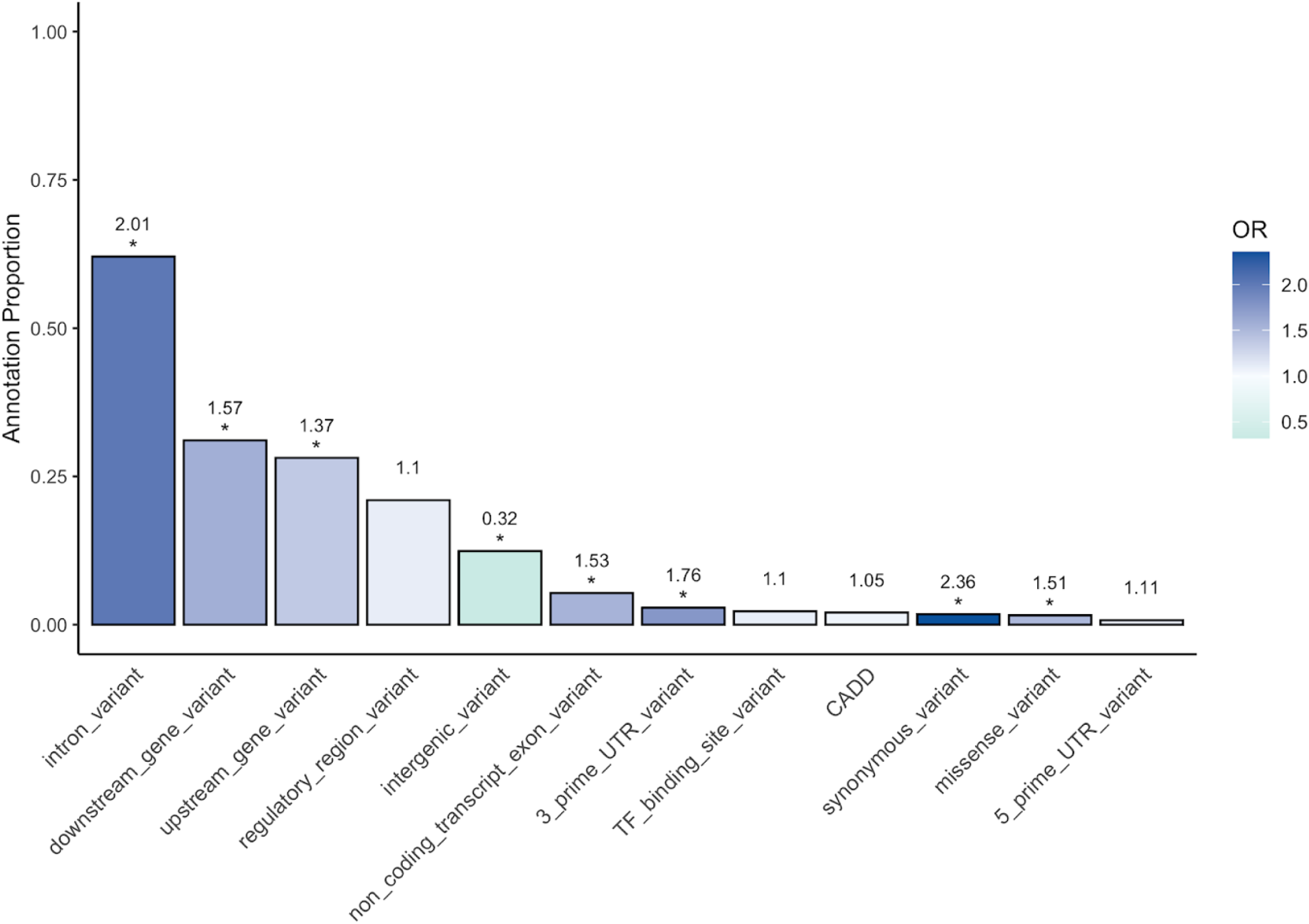
The enrichment and depletion of functional consequence annotations within the credible causal variants compared to the mapping region identified 8 significant annotations. The y-axis represents the proportion of variants within the credible causal variants that can be given that annotation. The annotation proportions do not add up to 1 because one variant can have multiple annotations. The colour of the bars represents the odds ratio (OR) from a Fisher’s exact test comparing counts of annotations in the credible causal variants versus counts of annotations in the rest of the mapping region. The stars represent OR which are significantly different from 1.

MAGMA gene set analysis^15^ identified 25 Gene Ontology biological process (**Supplementary Table 5**) that were significantly enriched, after multiple testing correction (*P*<3.23×10^−6^), for LOAD-associated variants. Subsequent conditional gene set analyses confirmed independent association of four out of these 25 gene-sets, reflecting the role of LOAD-associated genes in amyloid and tau plaque formation, protein catabolism of plaques, immune cell recruitment, and glial cells (**Supplementary Table 5**).

### Implicated Genes

Functional mapping of variants to genes based on position and expression quantitative trait loci (eQTL) information from brain and immune tissues/cells identified 989 genes mapped to one of the 38 genomic risk loci (**Supplementary Table 6**). Although the causal gene is not certain (see **Supplementary Results**), one of the positionally mapped genes (*ITGA2B*) within a novel locus (locus 28) is a potential drug target for LOAD treatment. The protein encoded by *ITGA2B* is a target for Abciximab, an antibody which inhibits platelet aggregation and is used to estimate concentrations of coated-platelets^17^. In patients with mild cognitive impairments, elevated coated-platelet levels are linked to increased risk of LOAD progression, and might constitute a relevant target for the mitigation of LOAD development.

Due to linkage disequilibrium (LD) and the inability to distinguish true causal variants from variants in LD, many of the mapped genes may be functionally irrelevant to LOAD. In order to highlight potentially relevant genes, eQTL data from immune tissues, brain, and microglia were colocalized with the genomic risk loci using Coloc^18^. We found 16 successful colocalizations for eight genes: *MADD, APH1B, GRN, AC004687*.*2, ACE, NTN5, CD33*, and *CASS4* (**Supplementary Table 7**). One notable borderline result was the colocalization of microglia eQTL data for *BIN1* with locus 4 with a posterior probability of 0.78. This result is notable because *BIN1* is a well-established LOAD risk gene^19^ and microglia appear to be enriched with expression of LOAD genes.

### Credible causal variants

Some loci were large and contained numerous genes, complicating identification of the causal variants and genes. Statistical fine-mapping with FINEMAP^20^ and SusieR^21^ narrowed down the signal to a smaller number of variants with the greatest probability to explain the association signal. Across the 36 fine-mapped loci (*APOE* and *HLA-DRB1* (MHC) loci were excluded due to complex LD) there were 386,521 variants; after fine-mapping there were 3822 unique variants across the FINEMAP^20^ 95% confidence set and the SusieR^21^ credible sets (hereafter “credible causal variants”). The median number of credible causal variants per locus was 74 and the median number of variants per locus that were included in the fine-mapping analysis was 11046. This represents a large reduction in the number of variants within a region. Full results of the fine-mapping are available in **Supplementary Table 8**.

### Active chromatin enrichment

All the variants included in the fine-mapping analyses (hereafter “mapping region”) were annotated as being in active or inactive chromatin across 127 cell types based on the ROADMAP Core 15-state model^22^. In all cell types, the credible causal variants were significantly enriched in active chromatin compared to the other variants included in the mapping region (**Supplementary Table 9**). As these analyses make comparisons amongst variants within genomic risk loci, which were shown to be enriched in active chromatin regions (**Supplementary Results**), this shows that credible causal variants harbor even greater potential for functional consequences. Cell types with credible causal variants most enriched for active chromatin included immune cells, brain tissue, and induced pluripotent cells derived from fibroblasts (iPS DF 19.11) **(Supplementary Figure 5**). The inclusion of immune cells in the top enriched cell types supports the findings from the genomic risk loci enrichment and again highlights immune genetics as important for LOAD.

### eQTL enrichment

The variants within the mapping region were annotated with eQTL information from 46 GTEx v8 tissues. All tissue types were significantly enriched with eQTLs in credible causal variants compared to the mapping region (ORs 3.70-28.60) (**Supplementary Table 10**). The large proportion and enrichment of eQTLs across all tissues in the credible causal variants compared to the mapping region highlights eQTLs as possible agents in driving the association signal. The pattern of enrichment did not appear to highlight any specific tissues of interest; rather it reflected the proportion of eQTLs within the credible causal variants. The cell types with the highest proportion of eQTLs in the credible causal variants tend to have the lowest enrichments and vice versa.

### Functional consequence enrichment

The variants within the mapping region were annotated for functional consequence as well as an estimate of deleteriousness (CADD score^23^) to determine whether the credible causal variants were enriched for an effect on genes (**Supplementary Table 11**). All annotations which imply proximity to genes were significantly enriched, except TF binding site and 5’ UTR (**Figure 2**). The only annotation which implies distance to genes (intergenic variants) was significantly depleted (OR=0.32, Prop=0.12, *P*_*Bonferroni*_=7.54×10^−152^). The most enriched variant annotation was synonymous variants (OR=2.36, Prop=0.018, *P*_*Bonferroni*_=1.14×10^−8^). Missense variants were slightly enriched (OR=1.51, Prop=0.016, *P*_*Bonferroni*_=0.037) and variants with CADD score > 15 were not significantly enriched. Of the 54 significant missense variants within the genomic risk loci, 29 were included in the fine-mapping region, however only 9 were identified in the fine-mapping credible sets. The relative lack of missense variant and CADD score (>15) enrichment combined with the high proportion and enrichment of eQTLs within the credible causal variants suggests that the association signal in the current study is largely driven through gene expression modulation rather than protein coding changes. This could be due to a true causal effect of eQTLs on LOAD pathology or due our focus on common variants which are less likely to be highly deleterious missense variants.

## Discussion

We performed the largest GWAS for LOAD to date, including 1,126,563 individuals, and identified 38 LOAD-associated loci, including seven novel loci. The data included both clinical cases and proxy cases, defined based on parental LOAD status, a strategy that was validated previously by us^8^ and others^24^. Through gene set analysis, tissue and single cell specificity analysis, colocalization, fine-mapping, and enrichment analyses, this study highlighted novel biological routes that connect genetic variants to LOAD pathology. These functional analyses all implicated immune cells and microglia as cells of interest which provided genetic support to the current understanding of LOAD pathology^25^. The seven novel loci were functionally annotated and fine-mapped to narrow down candidate causal genes (**Supplementary Results**). The functions of these candidate genes broadly align with the categories identified in the gene set analysis, with the addition of brain function, adult neurogenesis, protein-lipid interactions, and lipid efflux. Two of the novel loci have been previously associated with frontotemporal dementia (FTD)^26^. This signal is not driven by the non-medically verified LOAD cases in the UKB proxy LOAD data (**Supplementary Results**), which suggests that this region is pleiotropic for FTD or contains separate causal variants within the same LD blocks. In one of these novel loci (locus 28), *ITGA2B* mapped to the LOAD-associated region based on six significant variants in high LD (R^2^>0.9) with the lead variant in locus 28 (rs708382). One of the significant variants within *ITGA2B* was a high CADD score (19.8) exonic variant (rs5911). *ITGA2B* is a target of an existing drug (Abciximab) and may be a potential target for drug repurposing. However, more evidence supports *GRN* as the causal gene for locus 28; the association signal colocalized with an eQTL for *GRN* in brain tissue and *GRN* is a known FTD gene^27^. Further replication and fine-mapping will be beneficial in understanding which gene is driving the association in locus 28. For all LOAD-associated loci, we used genomic position and eQTLs in relevant tissues to map variants to genes and we then identified drugs which target those genes **(Supplementary Table 6**). Such work can form the basis for identification of novel drug targets, supporting the efforts from the pharmaceutical industry to develop effective LOAD treatment.

Future work focusing on fine-mapping, generating larger QTL databases in more specific cells types, and incorporating other ancestries will improve the interpretability of associated loci. Our fine-mapping was able to successfully narrow down associated loci to sets of causal variants; however, regional interpretation was limited due to the reliance on a well matched but not perfectly matched reference panel. Our colocalisation analysis identified a candidate causal gene in 8 of the 38 loci and we expect that larger and more specific QTL datasets will improve the number of successful colocalization. Yao *et al*. (2020)^28^ highlighted a need for higher sample size eQTL discovery and suggested that genes with smaller effect eQTLs are more likely to be causal for common traits. The identification of microglia, but not bulk brain tissue, as a cell/tissue type of interest in this study supported a finding in a recent single-cell epigenomic study^29^, which determined that investigating individual cell types will be more fruitful than bulk brain tissue for understanding the route from variant to LOAD pathology.

One important goal for LOAD GWAS is the identification of medically actionable information that can help in diagnosis or treatment in all populations. This study was limited in the ability to identify causal genes and in the applicability to non-European populations. Further study in non-European populations will improve the equity of genetic information and also help with fine-mapping of associated regions. Larger sample sizes of GWAS, epigenomic studies, and eQTL studies in all populations will improve identification and explanation of novel LOAD loci while increasing the applicability of these findings to a larger group of individuals. This could be accomplished by a push for facilitating data-sharing and global collaboration within the field of Alzheimer’s disease genetics. The current work provided genetic support for the role of immune cells, microglia, and eQTLs in LOAD, identified novel LOAD-associated regions, and highlighted the importance of collaboration to discern the biological process that mediate LOAD pathology.

### URLs

UK Biobank, http://ukbiobank.ac.uk; FUMA software, http://fuma.ctglab.nl; MAGMA software, http://ctg.cncr.nl/software/magma; mvGWAMA and effective sample size calculation, https://github.com/Kyoko-wtnb/mvGWAMA; LD Score Regression software, https://github.com/bulik/ldsc; LD Hub (GWAS summary statistics), http://ldsc.broadinstitute.org/; LD scores, https://data.broadinstitute.org/alkesgroup/LDSCORE/; NHGRI GWAS catalog, https://www.ebi.ac.uk/gwas/.

## Supporting information

Supplementary Results, Figures, and Methods

## Data Availability

Access to raw data can be requested via the Psychiatric Genomics Data Access portal https://www.med.unc.edu/pgc/shared-methods/open-source-philosophy/), UKBiobank, or 23andme. Summary statistics excluding 23andMe will be made available upon publication from https://ctg.cncr.nl/software/summary_statistics. Access to the full set including 23andme results can be obtained upon completion of a Data Transfer Agreement that protects the privacy of 23andMe participants. Please visit https://research.23andme.com/dataset-access/ to initiate a request. Summary statistics of the primary microglia eQTLs are also available from EGA (Accession ID: EGAD00001005736).

## Acknowledgements

We thank the research participants from 23andMe who made this study possible. Members of the 23andMe Research Team are: Michelle Agee, Stella Aslibekyan, Elizabeth Babalola, Robert K. Bell, Jessica Bielenberg, Katarzyna Bryc, Emily Bullis, Briana Cameron, Daniella Coker, Gabriel Cuellar Partida, Devika Dhamija, Sayantan Das, Sarah L. Elson, Teresa Filshtein, Kipper Fletez-Brant, Pierre Fontanillas, Will Freyman, Pooja M. Gandhi, Barry Hicks, David A. Hinds, Karen E. Huber, Ethan M. Jewett, Yunxuan Jiang, Aaron Kleinman, Katelyn Kukar, Vanessa Lane, Keng-Han Lin, Maya Lowe, Marie K. Luff, Jennifer C. McCreight, Matthew H. McIntyre, Kimberly F. McManus, Steven J. Micheletti, Meghan E. Moreno, Joanna L. Mountain, Sahar V. Mozaffari, Priyanka Nandakumar, Elizabeth S. Noblin, Jared O’Connell, Aaron A. Petrakovitz, G. David Poznik, Morgan Schumacher, Anjali J. Shastri, Janie F. Shelton, Jingchunzi Shi, Suyash Shringarpure, Chao Tian, Vinh Tran, Joyce Y. Tung, Xin Wang, Wei Wang, Catherine H. Weldon, and Peter Wilton. IGAP:We thank the International Genomics of Alzheimer’s Project (IGAP) for providing summary results data for these analyses. The investigators within IGAP contributed to the design and implementation of IGAP and/or provided data but did not participate in analysis or writing of this report. IGAP was made possible by the generous participation of the control subjects, the patients, and their families. The i–Select chips was funded by the French National Foundation on Alzheimer’s disease and related disorders. EADI was supported by the LABEX (laboratory of excellence program investment for the future) DISTALZ grant, Inserm, Institut Pasteur de Lille, Université de Lille 2 and the Lille University Hospital. GERAD/PERADES was supported by the Medical Research Council (Grant n° 503480), Alzheimer’s Research UK (Grant n° 503176), the Wellcome Trust (Grant n° 082604/2/07/Z) and German Federal Ministry of Education and Research (BMBF): Competence Network Dementia (CND) grant n° 01GI0102, 01GI0711, 01GI0420. CHARGE was partly supported by the NIH/NIA grant R01 AG033193 and the NIA AG081220 and AGES contract N01–AG– 12100, the NHLBI grant R01 HL105756, the Icelandic Heart Association, and the Erasmus Medical Center and Erasmus University. ADGC was supported by the NIH/NIA grants: U01 AG032984, U24 AG021886, U01 AG016976, and the Alzheimer’s Association grant ADGC– 10–196728.

HUNT: The Nord-Trøndelag Health Study (The HUNT Study) is a collaboration between HUNT Research Centre (Faculty of Medicine and Health Sciences, NTNU, Norwegian University of Science and Technology), Trøndelag County Council, Central Norway Regional Health Authority, and the Norwegian Institute of Public Health. The genotyping was financed by the National Institute of health (NIH), University of Michigan, The Norwegian Research council, and Central Norway Regional Health Authority and the Faculty of Medicine and Health Sciences, Norwegian University of Science and Technology (NTNU). The genotype quality control and imputation has been conducted by the K.G. Jebsen center for genetic epidemiology, Department of public health and nursing, Faculty of medicine and health sciences, Norwegian University of Science and Technology (NTNU).

The authors would like to thank the Norwegian Dementia Genetics Network (DemGene).This work was supported by the Research Council of Norway (RCN; 248980, 248778, 223273), Norwegian Regional Health Authorities, Norwegian Health Association and EU JPND: PMI-AD. This work was supported by the National Institutes of Health, National Institute on Aging R01 AG08724, R01 AG17561, R01 AG028555, and R01 AG060470.

Thank you to all the participants included in this study including the participants from Finngen, GR@CE, IGAP, UKB, DemGene, TwinGene, STSA, Gothenburg, ANMmerge, BioVU, 23andMe, HUNT, and deCODE.

AZ was supported by the Swedish Alzheimer Foundation (AF-930582, AF-646061, AF-741361), Stiftelsen Demensfonden, Magnus Bergvalls Stiftelse, Stiftelsen för Gamla Tjänarinnor, and Stiftelsen Handlanden Hjalmar Svenssons Forskningsfond. IS was supported by the Swedish state under the agreement between the Swedish government and the county councils, the ALF-agreement (ALF 716681), the Swedish Research Council (no 11267, 825-2012-5041, 2013-8717, 2015-02830, 2017-00639, 2019-01096), Swedish Research Council for Health, Working Life and Welfare (no 2001-2646, 2001-2835, 2001-2849, 2003-0234, 2004-0150, 2005-0762, 2006-0020, 2008-1229, 2008-1210, 2012-1138, 2004-0145, 2006-0596, 2008-1111, 2010-0870, 2013-1202, 2013-2300, 2013-2496), Swedish Brain Power, Hjärnfonden, Sweden (FO2016-0214, FO2018-0214, FO2019-0163), the Alzheimer’s Association Zenith Award (ZEN-01-3151), the Alzheimer’s Association Stephanie B. Overstreet Scholars (IIRG-00-2159), the Alzheimer’s Association (IIRG-03-6168, IIRG-09-131338) and the Bank of Sweden Tercentenary Foundation. SK was supported by the Swedish state under the agreement between the Swedish government and the county councils, the ALF-agreement (ALFGBG-81392, ALF GBG-771071), the Swedish Alzheimer Foundation (AF-842471, AF-737641), and the Swedish Research Council (2019-02075). MW was supported by the Swedish Research Council 2016-01590. HZ is a Wallenberg Scholar supported by grants from the Swedish Research Council (2018-02532), the European Research Council (681712), Swedish State Support for Clinical Research (ALFGBG-720931), the Alzheimer Drug Discovery Foundation (ADDF), USA (201809-2016862), the European Union’s Horizon 2020 research and innovation programme under the Marie Skłodowska-Curie grant agreement No 860197 (MIRIADE), and the UK Dementia Research Institute at UCL. KB was supported by the Swedish Research Council (2017-00915) and the Swedish state under the agreement between the Swedish government and the County Councils, the ALF-agreement (ALFGBG-715986). IKK receives funding from Swedish Research Council for Health, Working Life and Welfare (2018-01201) and the National Institutes of Health R01AG060470. NLP receives funding from the National Institutes of Health Grants No. R01AG059329, R01AG060470. CAR receives funding from the National Institutes of Health (NIH) including RF1AG058068, R01AG060470, R01AG059329, R01AG050595, and R01AG046938.

This work was funded by The Netherlands Organization for Scientific Research (NWO VICI 453-14-005), NWO Gravitation: BRAINSCAPES: A Roadmap from Neurogenetics to Neurobiology (Grant No. 024.004.012), and a European Research Council advanced grant (Grant No, ERC-2018-AdG GWAS2FUNC 834057). The analyses were carried out on the Genetic Cluster Computer, which is financed by the Netherlands Scientific Organization (NWO: 480-05-003), by the VU University, Amsterdam, The Netherlands, and by the Dutch Brain Foundation, and is hosted by the Dutch National Computing and Networking Services SurfSARA.

## Author contributions

DP and OAA conceived of the study. DPW performed the analyses. IEJ, JES, DP and OAA supervised analyses. DPW wrote the first draft of the manuscript. All authors contributed data and/or analyzed data and all authors critically reviewed the paper.

## Online Methods

### Dataset Processing

#### Participants

The data from the participants in this study were obtained from freely available summary statistics (N=145,339) and from genotype level data (N=981,224). Seven new cohorts were obtained since our previous analysis^1^ (as well as an increased deCODE sample), these cohorts contain 12,968 new cases and 488,616 new controls. An overview of the cohorts is available in Supplementary Table 1. Full description of each dataset, the quality control (QC) procedures, and the analysis protocol are available in the Supplementary Methods. In short, each dataset underwent initial QC, imputation, logistic/linear regression, and post-regression QC of the summary statistics using EasyQC^2^. If necessary, the data were converted to build GRCh37 before QC using the UCSC LiftOver tool^3^. During post-regression QC, each dataset was matched to the HRC or 1KG reference panel and variants with absolute allele frequency differences > 0.2 compared to the reference panel were removed. Variants with an imputation quality score < 0.8, MAC < 6, N < 30, or absolute beta or SE > 10 were removed. Low minor allele frequency (MAF) variants were removed; low MAF^4^ was defined as 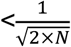.

#### Meta-analysis

All datasets were meta-analysed using mv-GWAMA, a sample size weighted method previously developed in Jansen *et al*. (2019)^1^. The option to account for overlapping individuals was not utilized because no datasets were expected to contain overlapping samples and the estimates of overlapping samples (genetic covariance intercepts) were unreliable due to low heritability of the datasets. The resulting Z-scores from the meta-analysis were converted to effect estimates (BETA) and standard errors (SE) using these formulae^5^:

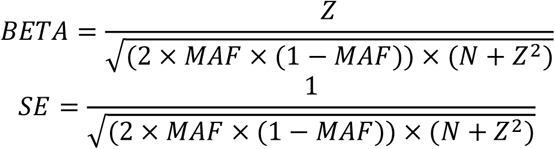

For a few (31) extremely significant *APOE* region variants, Z-scores could not be estimated so BETA and SE were calculated from a weighted average of the datasets which included those variants.

#### Genomic risk loci definition

We used FUMA v1.3.6^6^ to annotate and functionally map variants included in the meta-analysis. Genomic risk loci were defined around significant variants (<5×10^−8^); the genomic risk loci included all variants correlated (R^2^>0.6) with the most significant variant. The correlation estimates were defined using 1KG European reference information. The 1KG European reference panel was chosen over the UKB^7^ 10K reference panel because the meta-analysis included individuals from a range of European ancestries and this diversity would be better reflected in the 1KG European sample than the primarily British UKB sample. Genomic risk loci within 250Kb of each other are incorporated into the same locus.

Novel associated genomic risk loci are loci which do not overlap with variants identified as significant in previous studies of LOAD^1,8–14^. High and lower confidence novel loci are defined based on how well supported the lead variant is with other variants in LD. Variants with very few (<5) supporting variants are described as lower confidence. The edge of one of the novel loci (locus 34) was located within 2.7Mb of the edge of the *APOE* locus; in order to determine whether this was an independent locus GCTA-COJO^15^ was used to identify the independentlym associated SNPs within the *APOE* locus. The UKB participants were used as a reference panel to determine LD in the GCTA-COJO analysis. The independently associated SNPs were then conditioned on in an association analysis of the joint regions. The lead variant of locus 34 (rs2452170; GRCh37: 19:49213504) was still significant after conditioning on all independently associated variants within the *APOE* region (19:45000000-47000000).

#### Heritability and genetic correlation

Linkage disequilibrium score (LDSC) regression^16^ was used to estimate the heritability of the combined LOAD and proxy LOAD meta-analysis. LDSC^17^ was also used to determine the genetic correlation between a meta-analysis of datasets with clinically-diagnosed LOAD cases, and the UKB proxy LOAD dataset. Pre-calculated LD scores for LDSC were derived from the 1KG European reference population (https://data.broadinstitute.org/alkesgroup/LDSCORE/eur_w_ld_chr.tar.bz2). Heritability estimates were converted to a liability scale using the LOAD population prevalence of 0.05 and a sample prevalence of 0.0802. Heritability and genetic correlation estimates were calculated using HapMap3 variants only and the *APOE* region was excluded (GRCh37: 19:42353608-47713504). Further genetic correlations were determined using LDhub^18^, where all 855 traits were tested using the HapMap3 variants (http://ldsc.broadinstitute.org/static/media/w_hm3.noMHC.snplist.zip).

#### Gene-based and gene set analyses

Genome-wide gene association analysis was performed using MAGMA v1.08^19^. All variants in the GWAS outside of the MHC region (GRCh37: 6:28,477,797-33,448,354) that positionally map within one of the 19,019 protein coding genes were included to estimate the significance value of that gene. MAGMA gene set analysis was performed where variants map to 15,496 gene-sets from the MSigDB v7.0 database^20^. Forward selection of significantly associated gene-sets was performed using MAGMA v1.08 conditional analysis^21^. Initially the most significant gene-set was selected as a covariate and the remaining 24 gene-sets were analysed. The most significant gene-set from this conditional analysis was added as a covariate in addition to the previous gene-set and a new analysis was run. This process was repeated until no gene-set met the significance threshold (*P*<3.23×10^−6^). MAGMA tissue specificity analysis was performed in FUMA using 30 general tissue type gene expression profiles (from GTEx v8).

FUMA cell type specificity analysis^22^ utilises the MAGMA gene association results to identify cell types enriched in expression of trait associated genes. We focused on brain and immune related cell types with the inclusion of pancreas as a control, therefore selecting the following scRNA-seq datasets: Allen_Human_LGN_level1^23^, Allen_Human_LGN_level2^23^, Allen_Human_MTG_level1^23^, Allen_Human_MTG_level2^23^, DroNc_Human_Hippocampus^24^, DroNc_Mouse_Hippocampus^24^, GSE104276_Human_Prefrontal_cortex_all_ages^25^, GSE67835_Human_Cortex^26^, GSE81547_Human_Pancreas^27^, Linnarsson_GSE101601_Human_Temporal_cortex^28^, MouseCellAtlas_all^29^, PBMC_10x_68k^30^, and PsychENCODE_Adult^31^. Within-dataset conditional analyses results were reported to indicate which single cells are most likely to be disease relevant.

#### Gene mapping

The individual genomic risk loci were mapped to genes using FUMA v1.3.6^6^ using positional mapping and eQTL mapping. For positional mapping, all variants within 10Kb of a gene in the genomic risk locus were assigned to that gene. For eQTL mapping, variants were mapped to genes based on significant eQTL interactions in a collection of immune and brain tissues. Brain tissue eQTLs were used due to importance of brain tissue in LOAD pathology and immune tissue/cell eQTLs were used for gene mapping because MAGMA tissue specificity analysis highlighted immune tissues as tissues of interest. The brain and immune tissues eQTLs used for mapping were: Alasoo naive macrophage^32^, BLUEPRINT monocyte^33^, BLUEPRINT neutrophil^33^, BLUEPRINT T-cell^33^, BrainSeq Brain^34^, CEDAR B-cell^35^, CEDAR monocyte, CEDAR neutrophil^35^, CEDAR T-cell^35^, Fairfax B-cell^36^, Fairfax naive monocyte^37^, GENCORD T-cell^38^, Kasela CD4 T-cell^39^, Kasela CD8 T-cell^39^, Lepik Blood^40^, Naranbhai neutrophil^41^, Nedelec macrophage^42^, Quach monocyte^43^, Schwartzentruber sensory neuron^44^, TwinsUK blood^45^, PsychENCODE brain^31^, eQTLGen blood cis and trans^46^, BloodeQTL blood^47^, BIOS Blood^48^, xQTLServer blood^49^, CommonMind Consortium brain^50^, BRAINEAC brain^51^, GTEX v8 lymphocytes, brain, spleen, and whole blood. FUMA was also used to identify potential drug targets from the 989 mapped genes using Drugbank^52^.

#### Colocalization

All variants within 1.5Mb of the lead variant of each genomic risk loci were used in the colocalization analysis. The GWAS data and eQTL data were trimmed so that all variants overlap. Colocalization was performed per gene using coloc.abf from the Coloc R package^53^. Default priors were used for prior probability of association with the GWAS data and eQTL data. The prior probability of colocalization was set as 1×10^−6^ as recommended^54^. In all analyses BETA, SE^2^ (variance), case prevalence, and sample size from the GWAS data were used. In colocalization analyses with all tissues except microglia, nominal *P*, sample size, and minor allele frequency from the eQTL data were used. BETA, SE^2^ (variance), MAF, and sample size from the microglia eQTL data were used for colocalization. Colocalizations with a posterior probability > 0.8 were considered successful colocalizations. eQTL data from all tissues except microglia were obtained from the eQTL catalogue^55^. The microglia data was obtained from Young *et al*. (2019)^56^.

#### Fine-mapping

Fine-mapping was performed with SusieR v0.9.1^57^ and FINEMAP v1.4^58^ on all variants within 1.5Mb of the lead variant of each genomic risk loci. The *APOE* and *HLA-DRB1* (MHC) regions were excluded from fine-mapping due to the complicated LD structure. Chromosome 6 was excluded from fine-mapping because no associations outside of the HLA-DRB1 region existed and the two loci on chromosome 19 between 19:2550874-47713504 (GRCh37) were also excluded because they were within the *APOE* locus. Both SusieR and FINEMAP were used because it is recommended to use multiple fine-mapping methods in order to find consensus and increase the robustness of the results^59^. The sample size of the fine-mapping reference panel should be proportional to the sample size of the data being fine-mapped. A good-sized reference panel is 10% to 20% the sample size of the data^60^. UKB data was used as a reference panel for the fine-mapping because it had the largest sample size of the available reference panels and was the only available European reference panel to fulfill the criteria for a good-sized reference panel. In the SusieR analysis the reference panel was ∼10% the size of the GWAS data. In the FINEMAP analysis the reference panel was ∼40% the size of the GWAS data. The difference in size is due to FINEMAP allowing for individuals who have some genotypes missing to be included in the LD calculations from the reference panel. For FINEMAP, an LD matrix was generated using LDSTORE v2.0^60^ using 387,533 individuals included in the UKB. For SusieR, an LD matrix was generated using 100,000 individuals in R v3.4.3^61^. The 100,000 individuals were chosen for each locus as the top 100,000 people with the most genotyped variants in the locus in order to maintain the highest number of variants in the fine-mapping. Only the top 100,000 were chosen for computational feasibility and in order to maintain as many variants as possible while having a large reference panel. In both SusieR and FINEMAP, the meta-analysis data was trimmed to match the variants included in the LD reference. The maximum number of causal variants in the region was set to 10 in both methods. To generate the 95% FINEMAP confidence set, the variants included in the top N models in the configuration file were aggregated until the summed probability of the models reached 0.95. The SusieR 95% credible sets were generated by SusieR using default settings. The allele frequency in the UKB data and meta-analysis data of all the variants in the fine-mapping analyses were compared to identify outliers. No variants included in the confidence set or credible set had an allele frequency difference > 0.2.

#### Functional enrichment of significant association signals

All enrichment analyses were performed using a Fisher’s exact test (fisher.test) implemented in R 4.0.1^61^. For enrichment of genomic risk loci, all variants within the genomic risk loci were compared to all other variants present in the meta-analysis. For enrichment of the credible causal variants, all variants present in the 95% confidence set from FINEMAP and all variants in the credible set from successful SusieR analyses were compared to all other variants included in the mapping region.

Enrichment of active chromatin was performed using ROADMAP Core 15-state model annotation^62^ obtained from https://egg2.wustl.edu/roadmap/data/byFileType/chromhmmSegmentations/ChmmModels/coreMarks/jointModel/final/all.mnemonics.bedFiles.tgz. For each of the 127 cell types, all variants within the analysis were annotated with one of the 15 states using the R package Genomic Ranges^63^. All variants annotated with a state < 8 were defined as being within active chromatin. The enrichment of active chromatin within the specified region was performed for each of the cell types and the resulting *P*-values were corrected for 127 tests using Bonferroni correction.

Enrichment of eQTLs was performed using all tissues in the GTEx v8 dataset. Significant variant-gene pairs were downloaded from https://storage.googleapis.com/gtex_analysis_v8/single_tissue_qtl_data/GTEx_Analysis_v8_eQTL.tar. The significant eQTL variants were overlapped with the variants included in the enrichment analysis and eQTL counts within and outside the specified regions were generated for Fisher’s exact tests. The resulting *P*-values were corrected for the inclusion of 47 tissues from the GTEx v8 dataset using Bonferroni correction. The enrichment plots were generated using the R package ggplot2^64^.

Enrichment of functional consequences was performed using VEP^65^ and ANNOVAR^66^. All of the variants outside of the MHC region (GRCh37: 6:28,477,797-33,448,354) in the meta-analysis and the genomic risk loci variants were annotated using ANNOVAR and FASTA sequences for all annotated transcripts in RefSeq Gene^67^. The counts of each functional consequence in the genomic risk loci were compared to all variants outside of the loci. The significance threshold for the annotation enrichment was adjusted for the 11 tests performed (*P <* 0.0045). The mapping regions from the fine-mapping analysis were annotated using VEP to identify functional consequences and CADD score^68^ for exonic variants. The counts of each functional consequence in the credible causal variants were compared to the variants in the mapping region. Exonic variants were annotated with a CADD score and counts of credible causal variants with a CADD score >15 were compared to all other variants in the mapping region. The CADD score threshold was chosen because 15 was the median CADD score for the median value for all possible canonical splice site changes and non-synonymous variants in CADD v1.0. The significance threshold was adjusted for the 12 tests performed for the annotation enrichment (*P* < 0.0042).

