## Supplementary Results, Figures, and Methods for "Largest GWAS (N=1,126,563) of Alzheimer’s Disease Implicates Microglia and Immune Cells"

#### Author List and affiliations

Douglas P Wightman[1], Iris E Jansen[1], Jeanne E. Savage[1], Alexey A Shadrin[2], Shahram Bahrami[2–4], Arvid Rongve[5,6], Sigrid Børte[3,7,8], Bendik S Winsvold[8–10], Ole Kristian Drange[11,12], Amy E Martinsen[3,8,9], Anne Heidi Skogholt[8,13], Cristen Willer[14], Geir Bråthen[15–17], Ingunn Bosnes[11,18], Jonas Bille Nielsen[8,14,19], Lars Fritsche[20], Laurent F. Thomas[8,13], Linda M Pedersen[9], Maiken E Gabrielsen[8], Marianne Bakke Johnsen[3,7,8], Tore Wergeland Meisingset[15,16], Wei Zhou[21,22], Petra Proitsi[23], Angela Hodges[23], Richard Dobson[23–25], Latha Velayudhan[23], 23andMe Research Team[26], Julia M Sealock[27,28], Lea K Davis[27–29], Nancy L. Pedersen[30], Chandra A. Reynolds[31], Ida K. Karlsson[30,32], Sigurdur Magnusson[33], Hreinn Stefansson[33], Steinunn Thordardottir[34], Palmi V. Jonsson[34], Jon Snaedal[34], Anna Zettergren[35], Ingmar Skoog[35,36], Silke Kern[35,36], Margda Waern[35,37], Henrik Zetterberg[38–41], Kaj Blennow[40,41], Eystein Stordal[11,18], Kristian Hveem[8,42], John-Anker Zwart[3,8,9], Lavinia Athanasiu[2,4], Ingvild Saltvedt[15,17], Sigrid B Sando[15,16], Ingun Ulstein[43], Srdjan Djurovic[2], Tormod Fladby[3], Dag Aarsland[23,44,45], Geir Selbæk[3,43,46], Stephan Ripke[22,47,48], Kari Stefansson[33], Ole A. Andreassen[2,4,49], Danielle Posthuma[1,50*]

1. *Department of Complex Trait Genetics, Center for Neurogenomics and Cognitive Research, Amsterdam Neuroscience, VU University Amsterdam, The Netherlands.*
2. *NORMENT Centre, University of Oslo, Oslo, Norway.*
3. *Institute of Clinical Medicine, University of Oslo, Oslo, Norway.*
4. *Division of Mental Health and Addiction, Oslo University Hospital, Oslo, Norway.*
5. *Department of Research and Innovation, Helse Fonna, Haugesund Hospital, Haugesund, Norway.*
6. *The University of Bergen, Institute of Clinical Medicine (K1), Bergen Norway.*
7. *Research and Communication Unit for Musculoskeletal Health (FORMI), Department of Research, Innovation and Education, Division of Clinical Neuroscience, Oslo University Hospital, Oslo, Norway.*
8. *K. G. Jebsen Center for Genetic Epidemiology, Department of Public Health and Nursing, Faculty of Medicine and Health Sciences, Norwegian University of Science and Technology, Trondheim, Norway.*
9. *Department of Research, Innovation and Education, Division of Clinical Neuroscience, Oslo University Hospital, Oslo, Norway.*
10. *Department of Neurology, Oslo University Hospital, Oslo, Norway.*
11. *Department of Mental Health, Faculty of Medicine and Health Sciences, Norwegian University of Science and Technology, Trondheim, Norway.*
12. *Division of Mental Health Care, St. Olavs Hospital, Trondheim University Hospital, Trondheim, Norway.*
13. *Department of Clinical and Molecular Medicine, Norwegian University of Science and Technology, Trondheim, Norway.*
14. *Department of Internal Medicine, Division of Cardiovascular Medicine, University of Michigan, Ann Arbor, 48109, MI, USA.*
15. *Department of Neuromedicine and Movement Science, Norwegian University of Science and Technology, Trondheim, Norway.*
16. *Department of Neurology and Clinical Neurophysiology, University Hospital of Trondheim, Norway.*
17. *Department of Geriatrics, St. Olav's Hospital, Trondheim University Hospital, Norway.*

18. *Department of Psychiatry, Hospital Namsos, Nord-Trøndelag Health Trust, Namsos, Norway.*
19. *Department of Epidemiology Research, Statens Serum Institut, Copenhagen, Denmark.*
20. *Center for Statistical Genetics, Department of Biostatistics, University of Michigan, Ann Arbor, 48109, MI, USA.*
21. *Department of Computational Medicine and Bioinformatics, University of Michigan, Ann Arbor, MI, USA*.
22. *Analytic and Translational Genetics Unit, Massachusetts General Hospital, Boston, Massachusetts, USA.*
23. *Institute of Psychiatry, Psychology and Neurosciences, King's College London*.
24. *NIHR Biomedical Research Centre for Mental Health and Biomedical Research Unit for Dementia at 16 South London and Maudsley NHS Foundation, London, UK*.
25. *Farr Institute of Health Informatics Research, UCL Institute of Health Informatics*.
26. *23andMe, Inc., Mountain View, CA, USA*.
27. *Division of Genetic Medicine, Department of Medicine Vanderbilt University Medical Center Nashville, TN, 37232, USA*.
28. *Vanderbilt Genetics Institute, Vanderbilt University Medical Center, Nashville, TN, 37232, USA*.
29. *511-A Light Hall,Vanderbilt University Medical Center,2215 Garland Ave Nashville, TN 37232*.
30. *Department of Medical Epidemiology and Biostatistics, Karolinska Institutet, Stockholm, Sweden*.
31. *Department of Psychology, University of California-Riverside, Riverside, CA, USA*.
32. *Institute of Gerontology and Aging Research Network – Jönköping (ARN-J), School of Health and Welfare, Jönköping University, Jönköping, Sweden*.
33. *deCODE Genetics/Amgen, Sturlugata 8, IS-101, Reykjavik, Iceland*.
34. *Department of Geriatric Medicine, Landspitali University Hospital, Reykjavik, Iceland*.
35. *Neuropsychiatric Epidemiology Unit, Department of Psychiatry and Neurochemistry, Institute of Neuroscience and Physiology, the Sahlgrenska Academy, Centre for Ageing and Health (AGECAP) at the University of Gothenburg, Sweden.*
36. *Region Västra Götaland, Sahlgrenska University Hospital, Psychiatry, Cognition and Old Age Psychiatry Clinic, Gothenburg, Sweden*.
37. *Region Västra Götaland, Sahlgrenska University Hospital, Psychosis Clinic, Gothenburg, Sweden.*
38. *Department of Neurodegenerative Disease, UCL Institute of Neurology, London, United Kingdom.*
39. *UK Dementia Research Institute at UCL, London, United Kingdom.*
40. *Department of Psychiatry and Neurochemistry, Institute of Neuroscience and Physiology, the Sahlgrenska Academy at the University of Gothenburg, Mölndal, Sweden.*
41. *Clinical Neurochemistry Laboratory, Sahlgrenska University Hospital, Mölndal, Sweden.*
42. *HUNT Research Center, Department of Public Health and Nursing, Faculty of Medicine and Health Sciences, Norwegian University of Science and Technology, Trondheim, Norway.*
43. *Department of Geriatric Medicine, Oslo University Hospital, Oslo, Norway*.
44. *Centre of Age-Related Medicine, Stavanger University Hospital, Norway*.
45. *Institute of Psychiatry, Psychology & Neuroscience, PO 70, 16 De Crespigny Park, London, SE58AF*.
46. *Norwegian National Advisory Unit on Ageing and Health, Vestfold Hospital Trust, Tønsberg, Norway.*
47. Stanley Center for Psychiatric Research, Broad Institute of MIT and Harvard, Cambridge, MA, USA.
48. *Department of Psychiatry and Psychotherapy, Charité–Universitätsmedizin, Berlin,*

*Germany*.
49. *Oslo University Hospital, Kirkeveien 166, 0407 Oslo, Norway*.
50. *Department of Child and Adolescent Psychiatry and Pediatric Psychology, Section Complex Trait Genetics, Amsterdam Neuroscience, Vrije Universiteit Medical Center, Amsterdam University Medical Center, Amsterdam, The Netherlands*

*Correspondence should be addressed to: Danielle Posthuma: Department of Complex Trait Genetics, VU University, De Boelelaan 1085, 1081 HV, Amsterdam, The Netherlands. Phone: +31 20 598 2823, Fax: +31 20 5986926,

### Table of Contents

***Author List and affiliations*** ***1***
***Supplementary*** ***5***
**Supplementary Results** **5**
Independence of *NTN5* 5
Proxy vs case-control LOAD 5
Genomic risk loci enrichment 5
Active chromatin enrichment 5
Functional consequence enrichment 6
Novel Loci 6
Inconclusive Loci 7
Brain Regional Gene Expression 8
**Supplementary Figures** **8**
**Supplementary Methods** **20**
Datasets 20
deCODE 20
UK Biobank 20
Nord-Trøndelag Health Study (HUNT) 21
23andMe 21
BioVU 22
DemGene, TwinGene, STSA, Gothenburg, and ANMmerge 23
IGAP 24
Finngen 24
GR@CE 25
Brain regional gene expression 25
***References*** ***25***

### Supplementary

#### Supplementary Results

##### Independence of *NTN5*

One of the novel loci (*NTN5*) was within 2.7Mb of the *APOE* locus. As the signal of the *APOE* region is so strong, variants in low linkage disequilibrium (LD) with the region are likely to show a statistically significant signal even if there is no actual effect. To determine whether the *NTN5* locus is novel and independent of *APOE*, we used GCTA-COJO[1] to identify independent associated SNPs within the *APOE* locus (**see Online Methods**). The lead variant (rs2452170, GRCh37: 19:49213504) in the *NTN5* region showed no change in significance after conditioning on the associated *APOE* variants ($P$=1.72x$10^{-8}$, $P_{conditioned}$=1.60x$10^{-8}$). The lead variant in the *NTN5* region is in very low LD ($R^2$ < 0.007) with any of the independent associated SNPs in the *APOE* locus and in even lower LD ($R^2$=0.001) with the actual *APOE* variants (rs429358 and rs7412) in the European 1000 Genomes reference (1KG) population[2]. We thus conclude that the novel *NTN5* locus is independent from the APOE signal.

##### Proxy vs case-control LOAD

The genetic correlation between the proxy LOAD GWAS results and the case-control LOAD results was 0.83 (SE=0.21, $P$=6.61x$10^{-5}$) which is on par with the genetic correlation between proxy and case-control LOAD in our previous studies[3]. The high correlation suggests that the proxy design is a good estimate for LOAD status when the genotyped individual is too young to present the phenotype. However, there are differences between the results when specifying the phenotypes differently. Supplementary Figure 1 shows that the novel regions identified in the full meta-analysis do not have much significance in the proxy data alone. The *TMEM106B*, *GRN*, and *NTN5* regions did not have any variants with a P-value <0.0005 so none of the variants in that region were included in the Manhattan plot. Interestingly, *TMEM106B* and *GRN* are genes previously associated with frontotemporal dementia[4] and one would expect the LOAD proxy results to be driving this association due to the potential inclusion of dementia patients as cases but the association signal appears to be absent in proxy results. Supplementary Figure 2 shows that these genes do have relatively strong, albeit non-significant, signals in the results from the case-control data. Further exploration of the novel regions in an independent sample will be valuable in determining the role of these genes in LOAD.

##### Genomic risk loci enrichment

###### *Active chromatin enrichment*

The genomic risk loci (excluding the *HLA-DRB1* (MHC) region) contained 45,479 variants in total. An insight into the functional annotation of these variants may highlight potential routes from variant to phenotype. All the variants in the genome were annotated as being in active or inactive chromatin across 127 cell types based on the ROADMAP Core 15-state model[5] In all 127 cell types, the genomic risk loci variants were significantly enriched in variants within active chromatin compared to all variants included in the meta-analysis (**Supplementary Table 12**). The odds ratio (OR) of enrichment ranged from 4.34 to 1.71, with the top 5 cell types consisting of immune related cell types (**Supplementary**

**Figure 6**). The least enriched cell types were also significantly enriched compared to the rest of the genome, this is likely due to the genomic risk loci being located in gene dense regions which are likely to be more active in all cell types compared to the whole genome. The pattern of enrichment and proportion of active chromatin across the cell types again prioritizes immune cells as cell types of interest.

*Functional consequence enrichment*

The variants within and outside the genomic risk loci were also compared based on their functional consequence determined by ANNOVAR[6] (**Supplementary Figure 7; Supplementary Table 13**). The majority of the variants within the genomic risk loci are intronic and intergenic (prop=0.49, prop=0.36). The intergenic and ncRNA intronic variants were the only variant types to be significantly depleted in the genomic risk loci (OR=0.72, $P_{bonferroni}$=<1x10$^{-323}$; OR=0.76, $P_{bonferroni}$=1.20x10$^{-39}$), all other annotations, except ncRNA splicing variants, were significantly enriched. Splicing variants were the most enriched (OR=4.16, $P_{bonferroni}$=0.0098). These results suggest that the genomic risk loci are regions rich in genes, and that splicing may be an important mechanism through which effects of these genes on LOAD are regulated.

### Novel Loci

Locus 8 contains 71 variants in LD where the lead variant is rs6891966 (*P*=7.91x10$^{-10}$). This variant is located in an intron of *HAVCR2* (**Supplementary Figure 8**). *HAVCR1* and *TIMD4* also map to this region based on brain eQTLs (PsychENCODE). SusieR fine-mapping did not highlight any high posterior probability variants. FINEMAP identified 7 variants which have PIP>0.99, all 7 of these variants are *HAVCR2* intron variants. *HAVCR2* is preferentially expressed in aged microglia[7]. *HAVCR2* was included as one of the top 100 enriched transcripts in brain and microglia and was included in a cluster of transcripts which are involved in sensing endogenous ligands and microbes[8]. Havcr2 has been suggested to bind to phosphatidylserine on cell surfaces to mediate apoptosis[9]. Another study found Havcr2 to significantly interact with amyloid precursor protein[10]. *HAVCR2* expressed in microglia could mediate binding with cells to initiate apoptosis or binding to amyloid beta plaque. *TIMD4* is another gene in this region which shows similar function to *HAVCR2*, it binds to phosphatidylserine on cell surfaces to mediate apoptosis and microglia lacking *TIMD4* receptors have reduced apoptotic clearance[11].

Locus 12 and locus 28 have both been previously associated with dementia[4]. The lead variant in locus 12 was rs5011436, an intron variant in *TMEM106B* with *P*-value of 2.7x10$^{-9}$ (**Supplementary Figure 9**). Neither fine-mapping tools identified any variants with high PIP. However, a nearby variant (rs3173615) is an exonic variant with a CADD score of 21.2. This variant has been discussed as the association signal driving variant in frontotemporal dementia (FTD) by causing decreased *TMEM106B* protein abundance through increasing protein degradation[12]. The lead variant in locus 28 was rs708382, a downstream variant of *RPL7L1P5* with a *P*-value of 1.98x10$^{-9}$(**Supplementary Figure 10**). Close to the lead variant is an exonic variant (rs5911) in *ITGA2B* with a CADD score of 19.8. This variant, and the gene it exists within (*ITGA2B*), appears to impact platelets and cardiovascular disease. In this region, eQTLs for *GRN* in blood and brain colocalized with the GWAS signal. *GRN* is a known FTD gene[13].

Locus 34 contains 71 variants in LD, with a lead variant (rs2452170) between *FUT2* and *MAMSTR* with a *P*-value of 1.72x10$^{-8}$ (**Supplementary Figure 11**). This region maps to 16 genes, two of which include exonic variants with high CADD scores. *FUT2* contains rs601338, a variant with a CADD score of 52 which introduces a stop codon. *FUT2* also contains another stop codon-introducing variant (rs602662), which has a CADD score of

22.5. The former variant has been previously associated with gastroenteritis and Crohn's disease, and the latter was associated with B12 blood levels[14]. A Mendelian randomisation study of B12 levels and LOAD used rs602662 and other variants to determine that, opposed to observational analyses, B12 levels does not mediate LOAD pathology[15]. However, they used this variant under the assumption that it is not associated with LOAD and rs602662 was suggestively associated with LOAD in this meta-analysis (*P*-value=$5.12 \times 10^{-6}$). The other exonic variant (rs2287922) with a high CADD score (26.2) is located in *RASIP1*. *RASIP1* appears to be involved with maintenance of the blood brain barrier[16]. Blood brain barrier malfunction has been observed in AD[17]. In a recent unpublished study, *MAMSR* was identified as a gene which is differentially methylated in LOAD brains[18]. *NTN5* was implicated through colocalization of the GWAS signal with an eQTL in brain tissue. *NTN5* is highly expressed in neurogenic regions of the brain and is known to be involved in adult neurogenesis[19]. This locus is a difficult locus to interpret due to the range of genes and spread of association signal, however *NTN5* and *RASIP1* appear to have the most evidence to support their association with LOAD.

### Inconclusive Loci

The first inconclusive region identified in this study (locus 1) was identified due to a rare (MAF=0.0041) variant (rs113020870). This variant is a synonymous exon variant within *AGRN* (**Supplementary Figure 12**). This locus was categorised as inconclusive because the lead variant is only supported by 2 other variants in LD ($R^2$>0.1). The variant is present in 3 datasets (23andMe, Finngen, and UKB). The effect direction in all 3 is the same and the *P*-values in the UKB and 23andMe datasets are below $5 \times 10^{-4}$ (**Supplementary Figure 13**). The gene *AGRN* codes for agrin, a protein which influences the functioning of excitatory synapse and the blood brain barrier and has been previously suggested to be important to neurological diseases like LOAD[20]. Another study identified that *AGRN* expression influenced amyloid-beta homeostasis in mouse models of LOAD[21]. Amyloid plaques have been found to consist of significant amounts of heparan sulfate proteoglycans (HSPGs), a family of proteins which contain agrin[22]. The same study suggested that microglia expressed complement receptor 3 receptors bind to HSPGs and amyloid in plaques in the LOAD brain. Together, the previous literature and this GWAS result highlights the possibility that agrin makes up a proportion of amyloid plaques and rare mutations within *AGRN* could affect the binding efficacy and clearance of these plaques by microglia.

The second inconclusive region (locus 3) consists of two variants in LD (**Supplementary Figure 14**); rs115186657 is the only significant variant (*P*=$1.33 \times 10^{-8}$; MAF=0.0035). This locus is categorised as inconclusive because it contains only 2 variants in LD ($R^2$>0.1). This variant is included in 2 datasets (23andMe and UKB); the variant has the same effect direction in both datasets and has a *P*-value $<1 \times 10^{-4}$ in both datasets (**Supplementary Figure 15**). This region contains no prospective genes, nor do any genes map to the region based on eQTL data. The closest gene to the non-significant variant is *FHL2*, there is some evidence that this protein interacts with the protein of a LOAD associated gene (*PSEN2*)[23]. The fine-mapping results of this locus identified 2 variants, rs115186657 (PIP=0.88) and rs143254526 (PIP=0.04), the former is the lead variant and the latter is located in *NCK2*. A recent unpublished study of AD[24] identified a novel significant variant (rs143080277) in *NCK2*, this variant is ~130Kb away from the end of the genomic risk locus identified in this study. It is likely that the region identified in our study is separate to the *NCK2* region but due to the large fine-mapped region there is some signal from the *NCK2* being included in this locus. The variant identified as significant in Schwartzentruber et al. (2020)[24] has a *P*-value of $4.11 \times 10^{-7}$ in this study, which could explain why it has been included in the credible causal set but with a low PIP.

The third inconclusive region (locus 7) contains a single significant variant (rs871269, $P=1.37\times10^{-9}$) with a MAF of 0.34 (**Supplementary Figure 16**). This locus is inconclusive because the lead variant is not supported by any other variants in LD ($R^2>0.1$). The lead variant is included in all but one dataset and 13 out of 17 datasets have the same effect direction (**Supplementary Figure 17**). The *P*-value is fairly modest in all datasets with the lowest *P*-value of 0.00034 in the UKB data. This variant is located in an intron of *TNIP1* and maps to *GPX3*, *TNIP1*, and *SLC36A1* based on eQTLs within blood tissue. The fine-mapping results from FINEMAP and SusieR both highlighted the lead variant as the only variant with high posterior probability of inclusion (PIP>0.99 in both). *TNIP1* contributes to hyperinflammation and has been previously identified in autoimmune GWAS[25]. *TNIP1* was included in a transcription module regulated by *BCL3* in mouse microglia[26]. This module was implicated in prolonged exposure to inflammation and aging of microglia. *BCL3* is a gene significantly associated with LOAD after conditioning for *APOE*[27] and was observed as upregulated in the LOAD brain[28].

#### Brain Regional Gene Expression

The regional brain expression of the genes implicated by eQTL mapping was examined using GAMBA[29] (**Supplementary Figure 18**). The mean gene expression of the 329 mapped genes were compared to a random selection of 329 significantly brain expressed genes which resulted in 27 regions which differed significantly (**Supplementary Table 14**). The significantly different regions with the highest mean were the left thalamus proper, caudal anterior cingulate, insula, pallidum, and postcentral gyrus. These regions have a range of functions including somatosensory function, emotion, and memory[30–34]. However, when the regional mean gene expressions of the mapped genes were compared to a random selection of genes (not just brain expressed genes) there were no significant differences between the mean gene expression of any region (**Supplementary Table 14**). This result may reflect the lack of brain specificity of the mapped genes which supports the finding in this study that LOAD risk is mediated through immune related cells (microglia) and highlights the importance of narrowing down associated regions to individual causal genes.

### Supplementary Figures

*Supplementary Figure 1*: Manhattan plot of the UKB proxy LOAD data indicates low association of the novel regions (green) identified in the full meta-analysis. Only variants with a $P< 0.0005$ are displayed so novel regions with *P-values* larger than this are not observable. The *APOE* region cannot be fully observed because the y-axis is limited to the top variant in the second most significant locus, $-\log10(1\times10^{-60})$, in order to display the less significant variants. The red line represents genome wide significance ($5\times10^{-8}$)

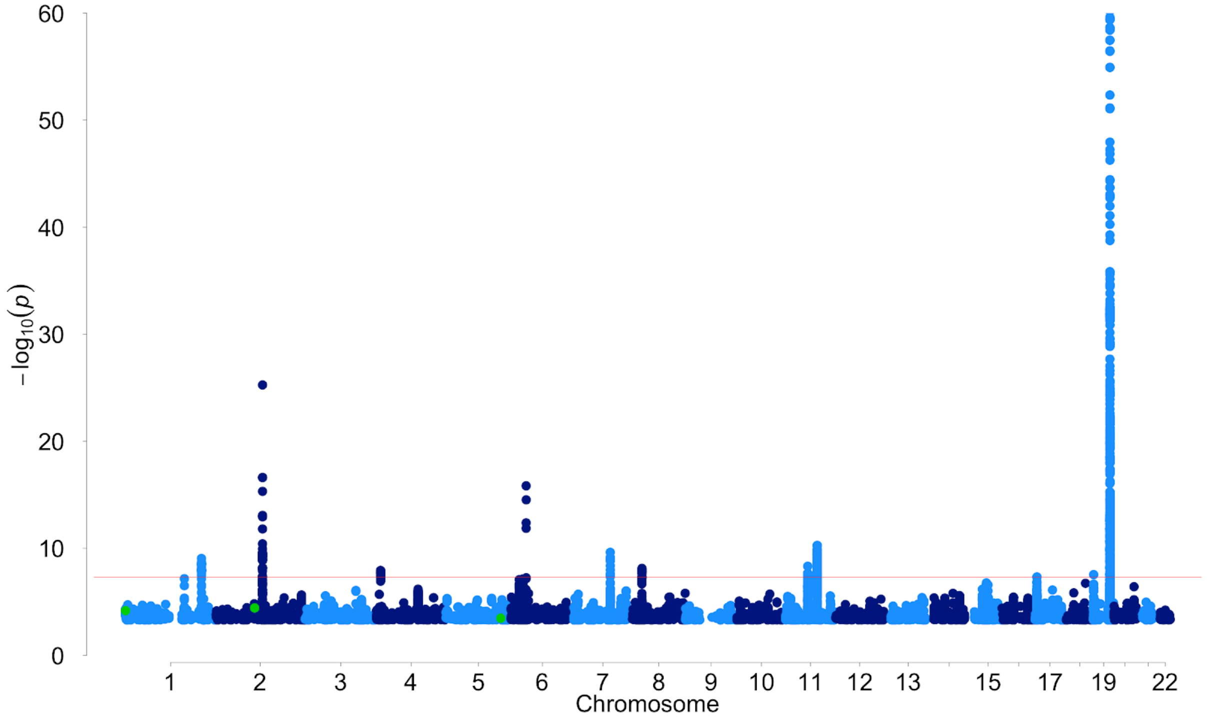


*Supplementary Figure 2*: Manhattan plot of the meta-analysis results of the LOAD case-control data indicates some association of identified in the full meta-analysis. Only variants with a *P*< 0.0005 are displayed. The *APOE* region cannot be fully observed because the y-axis is limited to the top variant in the second most significant locus, -log10($1x10^{-60}$), in order to display the less significant variants. The red line represents genome wide significance ($5x10^{-8}$)

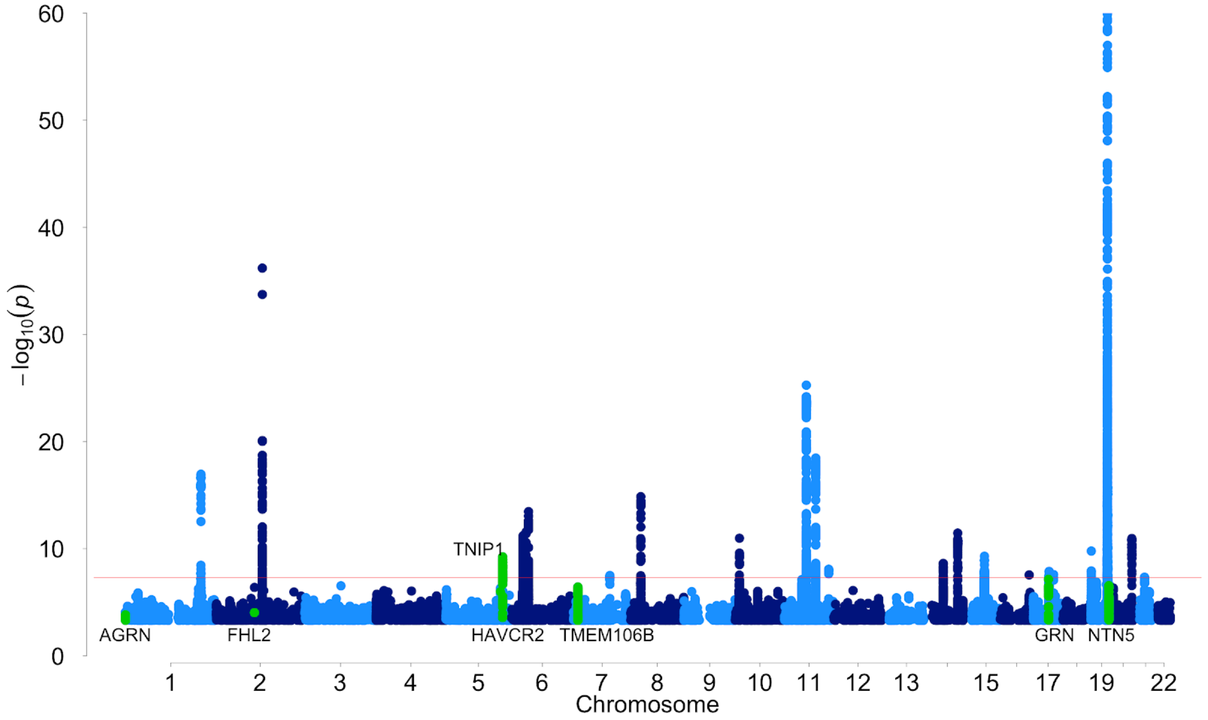

*Supplementary Figure 3*: MAGMA tissue specificity analysis identified spleen as a GTEx tissue with similar expression profiles to the LOAD gene association profile. The dotted line represents the significance threshold based on 30 tests. Significantly associated tissue is highlighted in dark blue. The full results and names of each tissue are available in **Supplementary Table 3**.

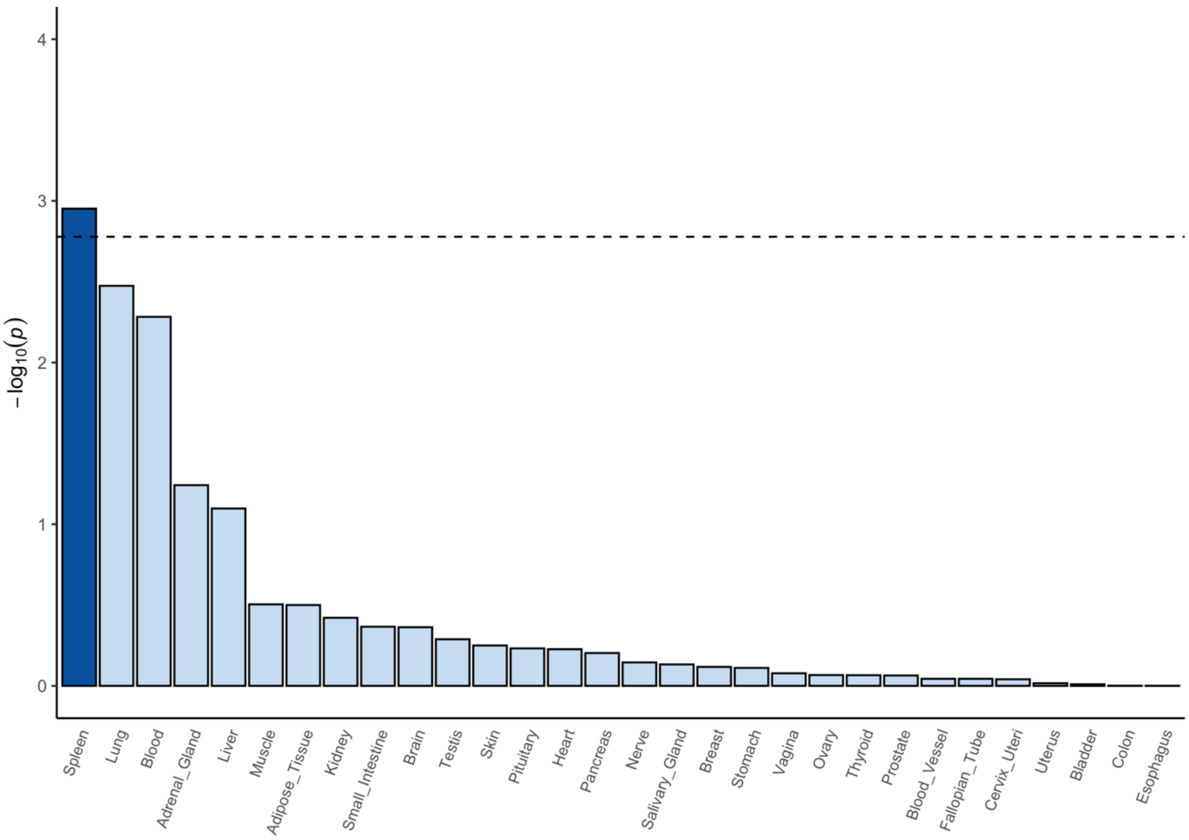


*Supplementary Figure 4*: Independent cell type associations based on within-dataset conditional analyses identifies microglia as the only cell type of interest. The cell specific expression profiles of microglia in 6 datasets are significantly associated ($P$<5.39x10$^{-5}$) with the LOAD association reflected in the MAGMA gene analysis. Microglia were significantly enriched in human lateral geniculate nucleus ($P$=1.11x10$^{-7}$), human middle temporal gyrus ($P$=6.41x10$^{-7}$), adult human brain ($P$=8.72x10$^{-6}$), mouse hippocampus ($P$=1.15x10$^{-5}$), human prefrontal cortex ($P$=1.28x10$^{-5}$), and brain macrophage (microglia) mouse brain ($P$=8.11x10$^{-6}$).

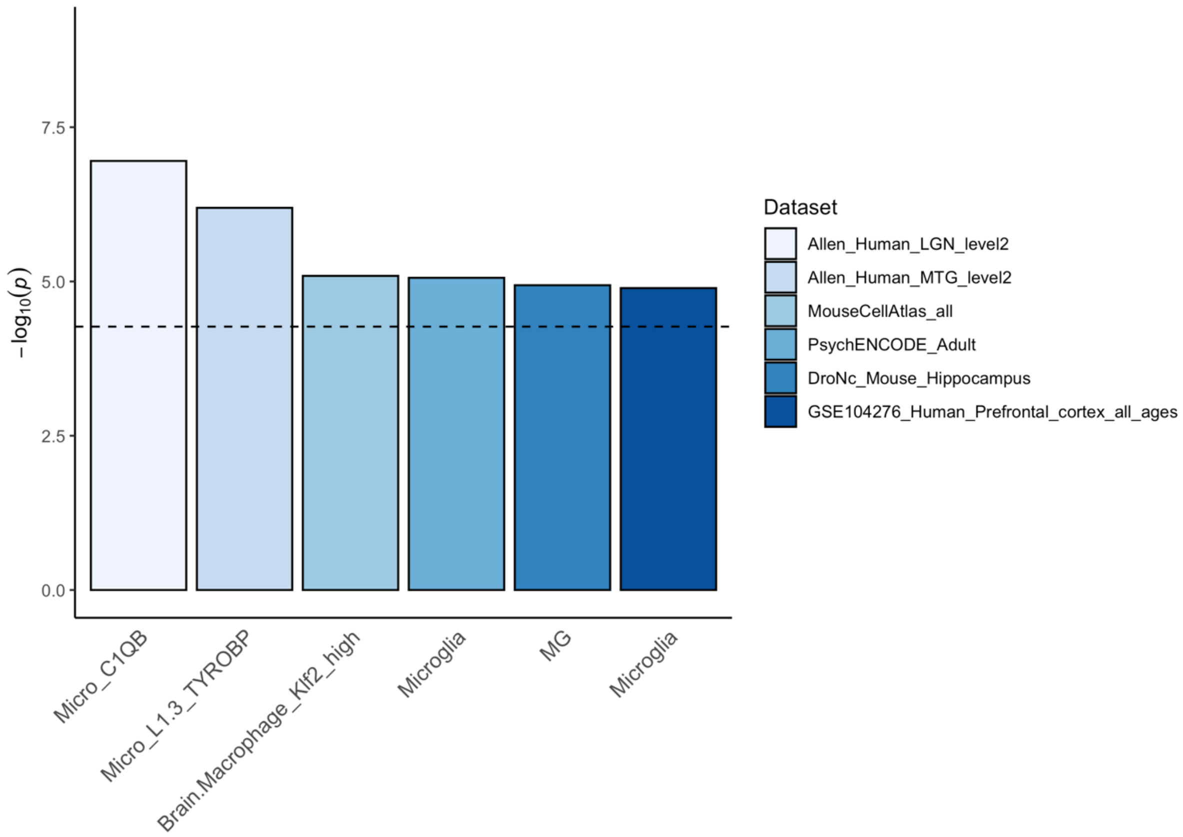


*Supplementary Figure 5*: The top 5 and bottom 5 cell types enriched for active chromatin in the credible causal variants compared to the mapping region highlights an induced pluripotent stem cell, brain tissue, and immune cells. The y-axis represents the proportion of variants within the credible causal variants which are in active chromatin for a given cell type. All cell types are significant after Bonferroni correction for 127 cell types. iPS_DF= induced pluripotent cells derived from fibroblasts (IPS DF 19.11). All OR were significantly different from 1.

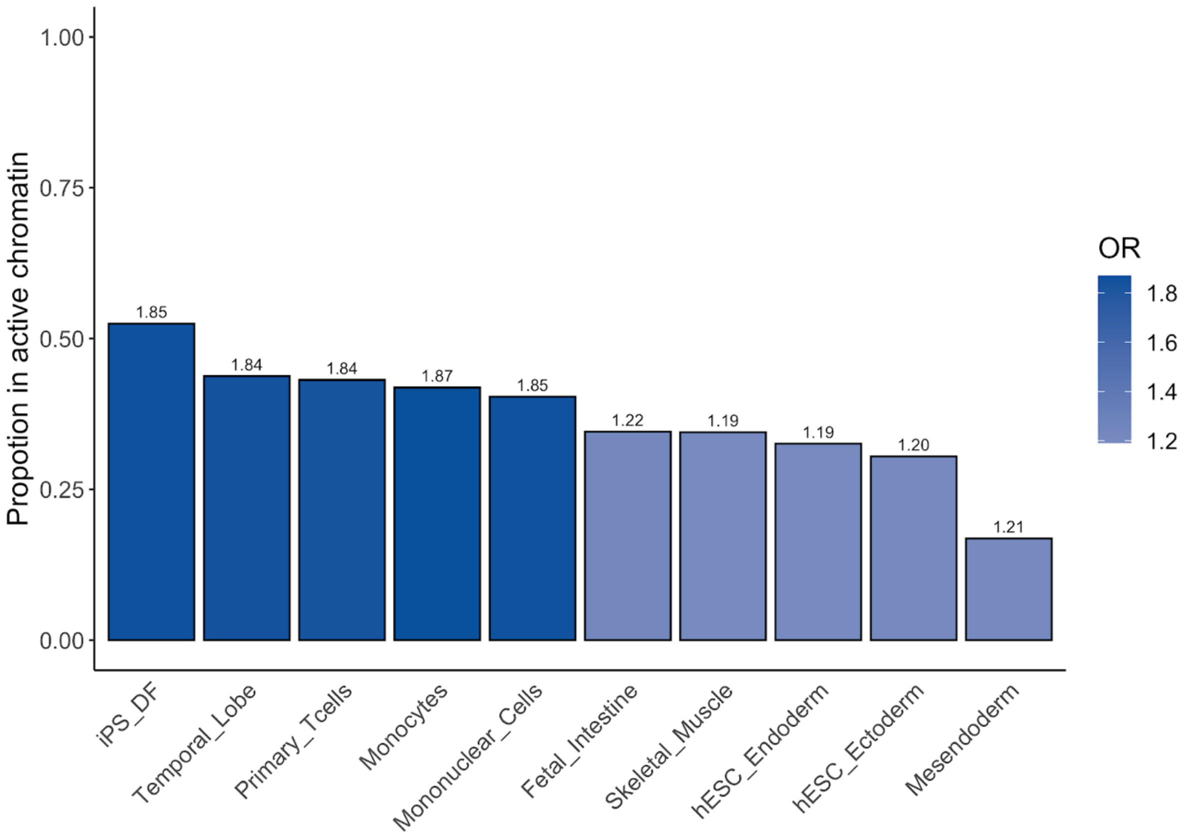


*Supplementary Figure 6*: The top 5 and bottom 5 cell types where active chromatin is most enriched in genomic risk loci compared to the rest of the genome highlights immune cells as enriched for active chromatin in LOAD regions of interest. The y-axis represents the proportion of the genomic risk locus that is in active chromatin in that cell type. The colour of the bars represents the odds ratio (OR) from a Fisher's exact test comparing counts of variants in active chromatin in the genomic risk loci vs counts of variants in active chromatin in the rest of the genome. All OR were significantly different from 1.

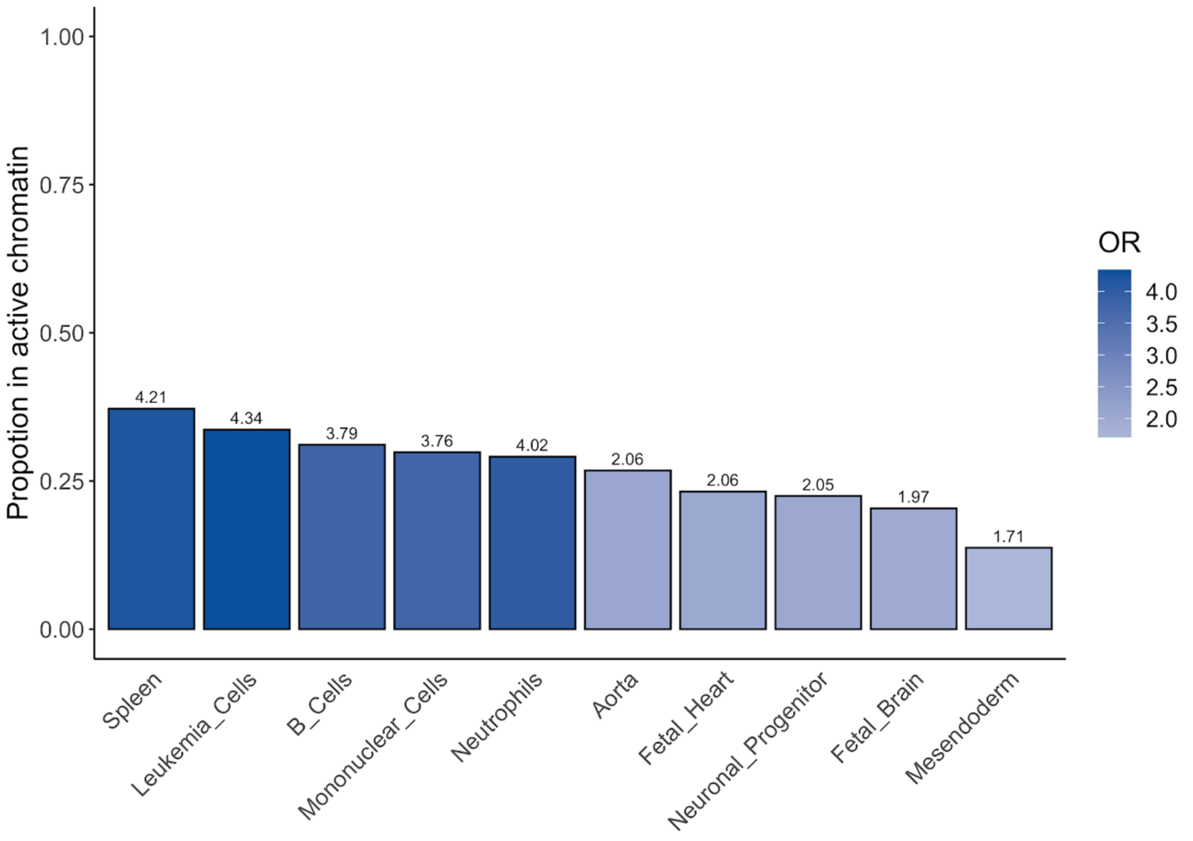


*Supplementary Figure 7*: ANNOVAR enrichment analyses identifies 10 significant differences between the number of annotations in the genomic risk loci compared to the rest of the genome. The y-axis represents the proportion of the genomic risk loci which falls into each annotation. The colour of the bars represents the odds ratio (OR) from a Fisher's exact test comparing counts of annotations in the genomic risk loci vs counts of annotations in the rest of the genome. The asterisks (*) represent OR which are significantly different from 1.

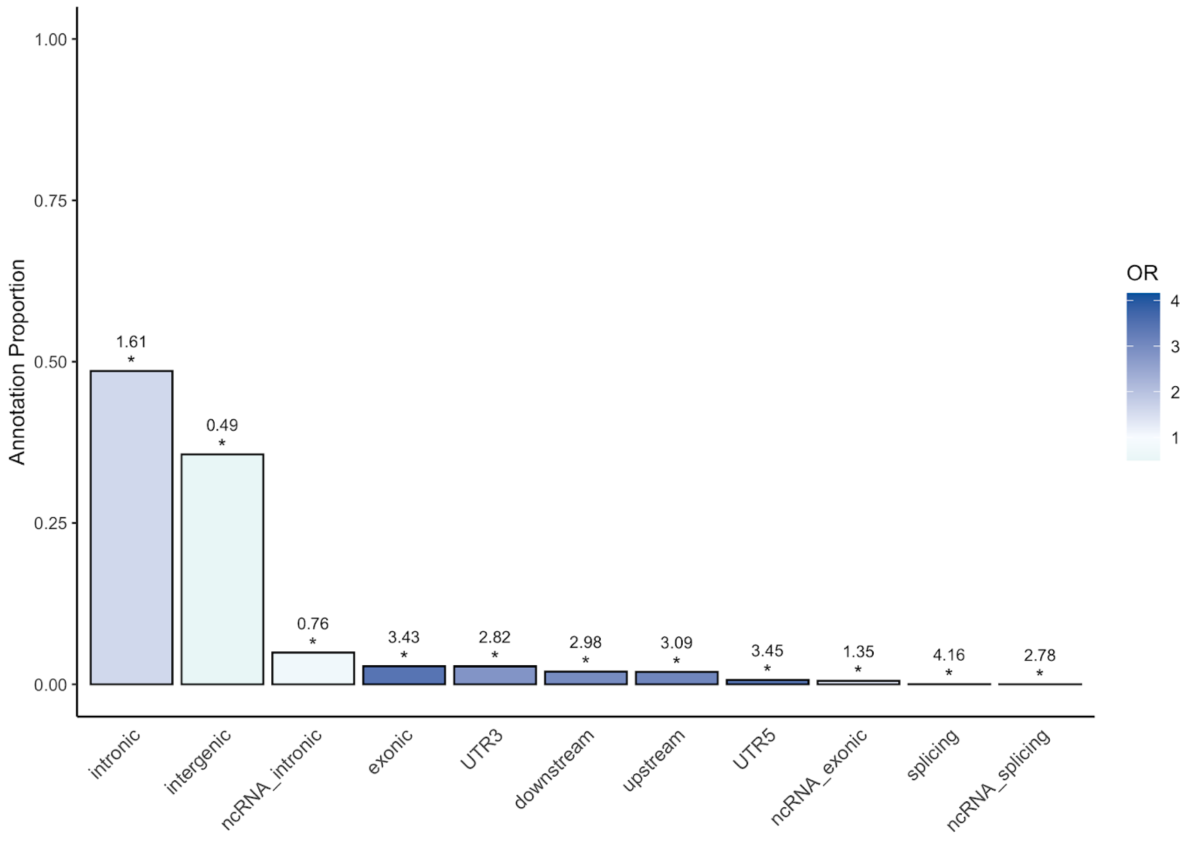


*Supplementary Figure 8*: Regional plot highlighting the lead variant of locus 8 and the genes of interest. The eQTLs and the genes which they map to are included below the regional Manhattan plot.

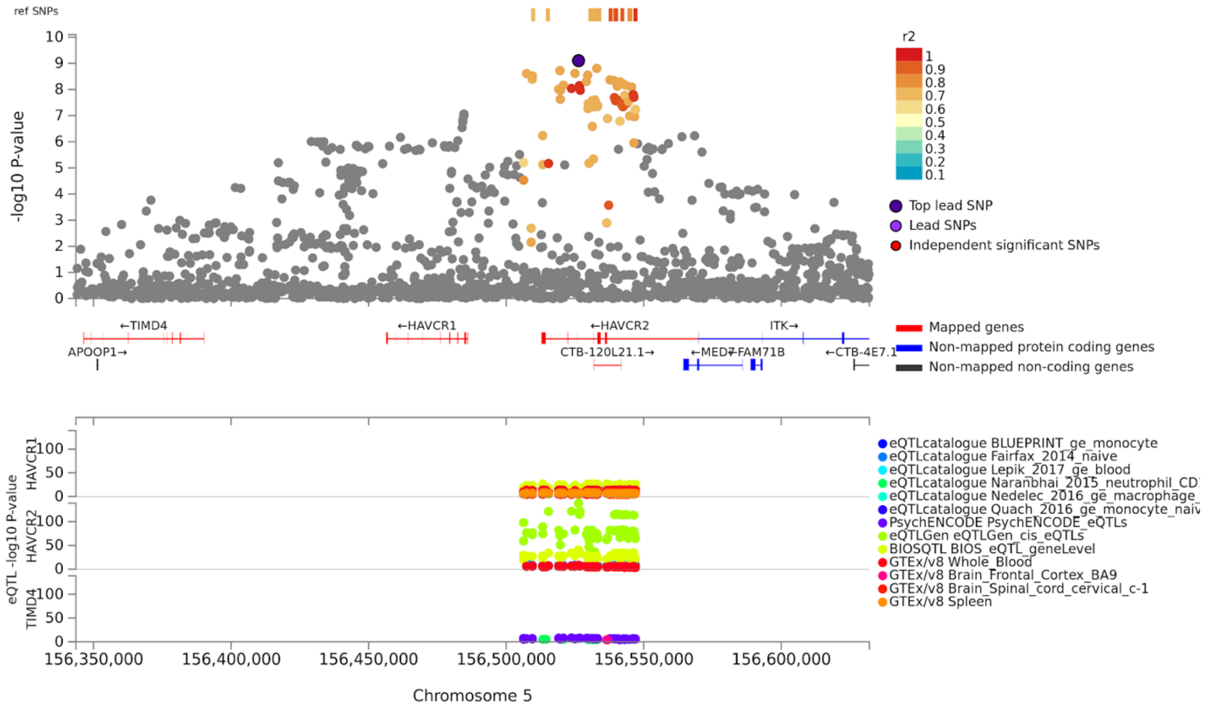

*Supplementary Figure 9*: Regional plot highlighting the lead variant of locus 12 and the gene of interest (*TMEM106B*). The CADD score of the included variants are included below the regional Manhattan plot.

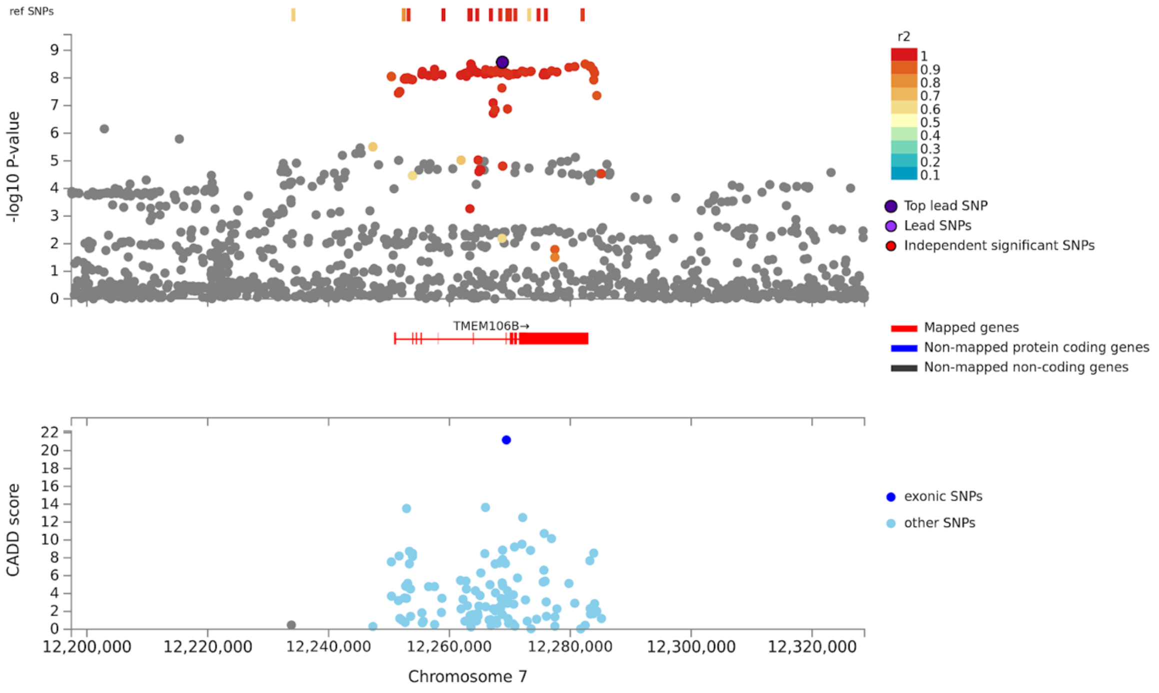


*Supplementary Figure 10*: Regional plot highlighting the lead variant of locus 28 and the genes of interest.

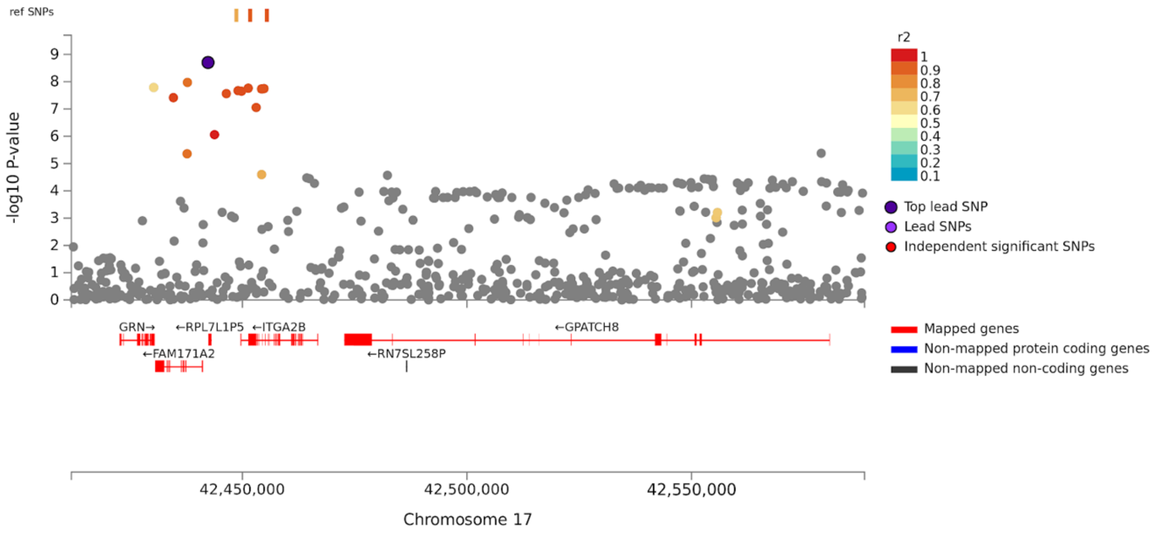

*Supplementary Figure 11*: Regional plot highlighting the lead variant of locus 34 and the genes of interest. The CADD score of the included variants are included below the regional Manhattan plot.

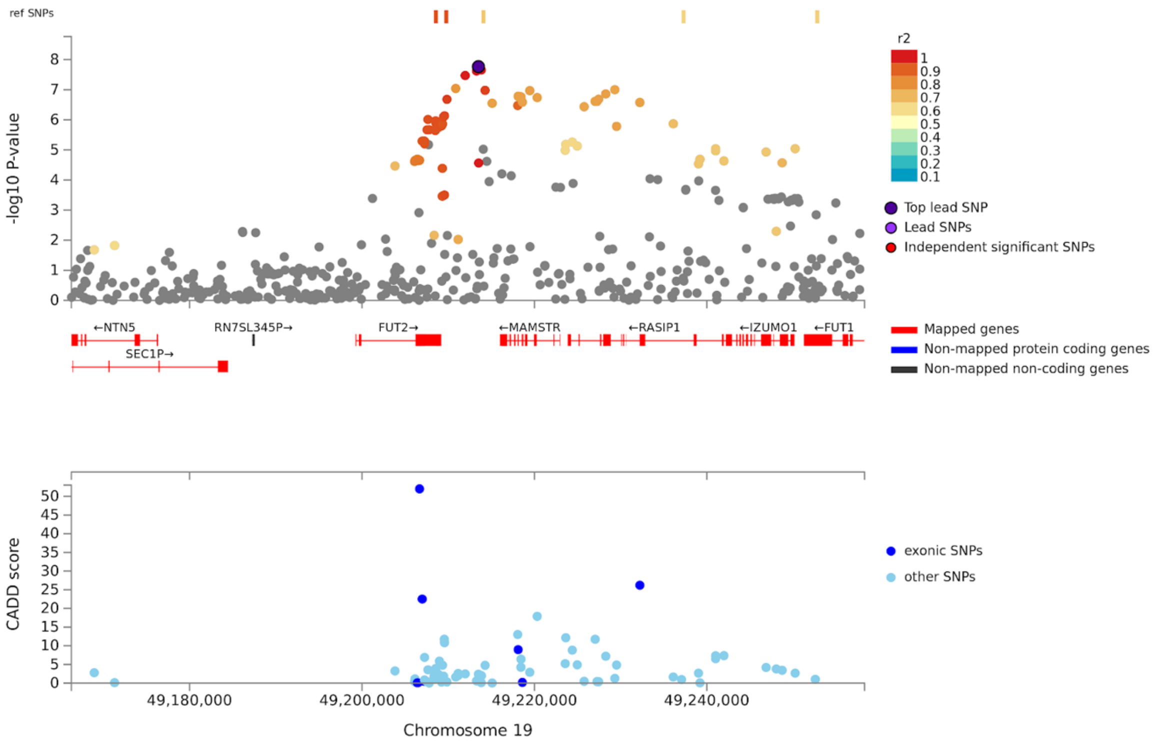


*Supplementary Figure 12*: Regional plot of locus 1 (inconclusive) indicating the lead variant and the two variants in LD in this
region.

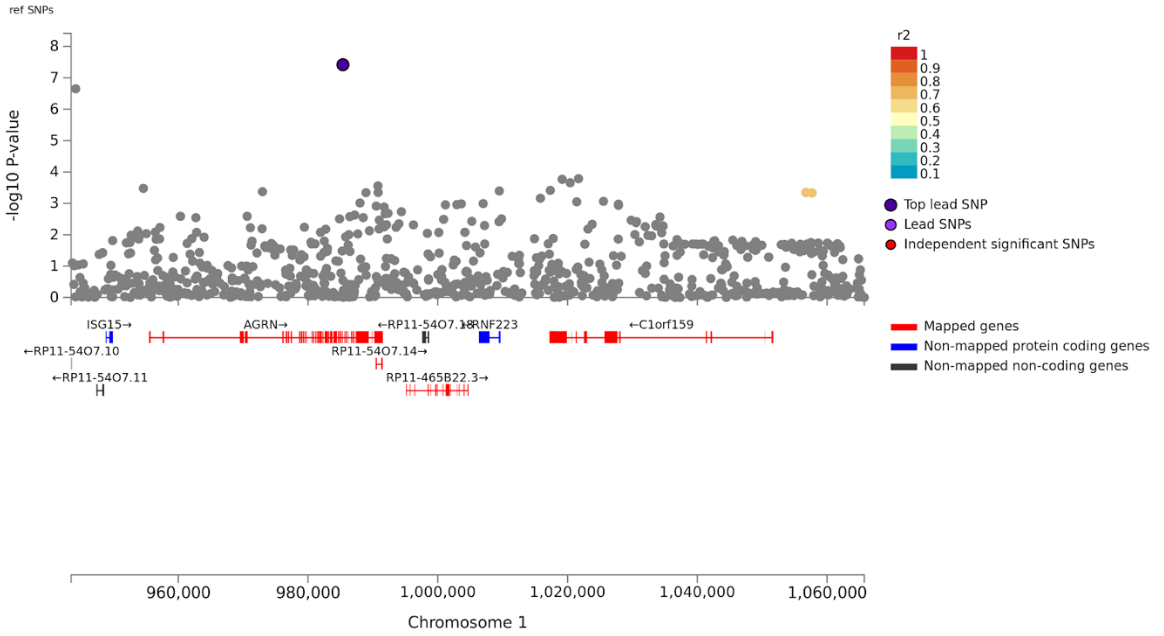

*Supplementary Figure 13*: Forest plot of rs113020870 highlighting the effect estimate (BETA), standard error (SE) and *P-value* of this variant in each dataset. The error bars represent 95% confidence intervals.

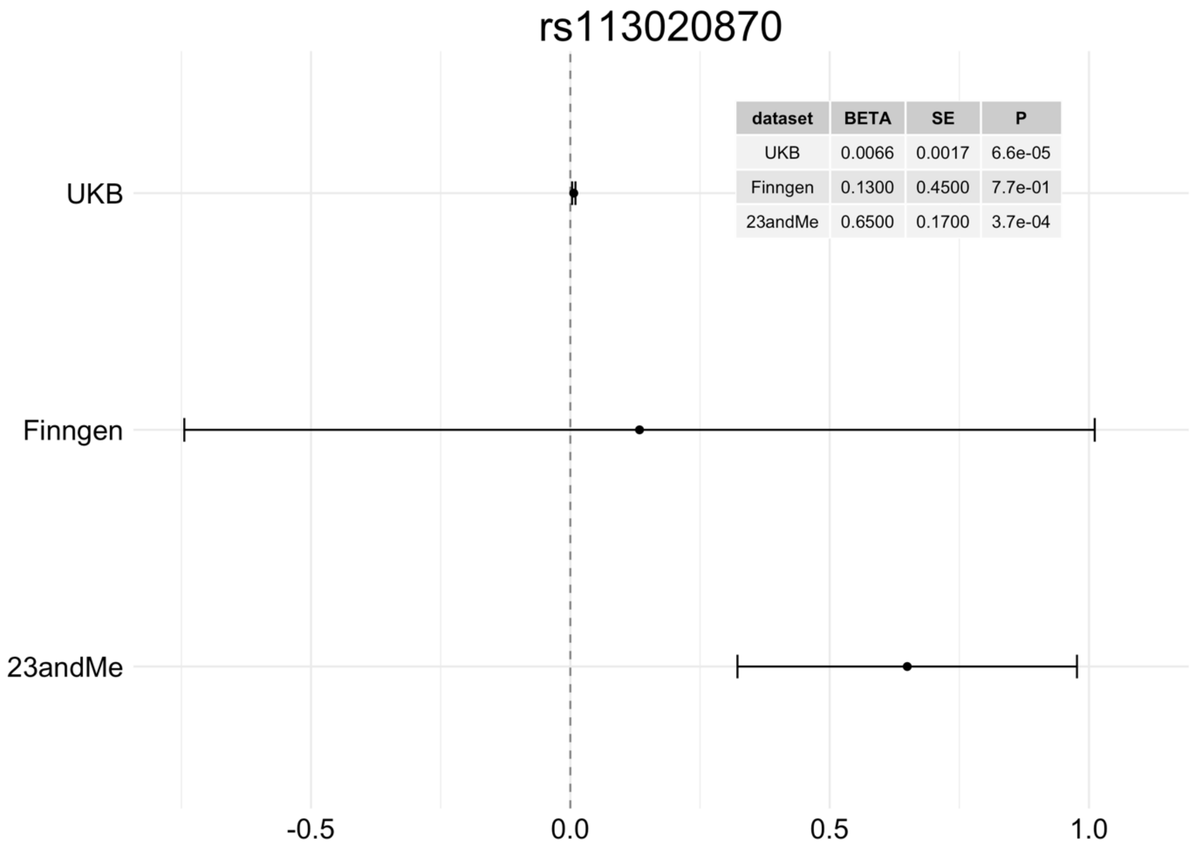


*Supplementary Figure 14*: Regional plot of locus 3 (inconclusive) highlighting the associated variant in the region and nearby genes.

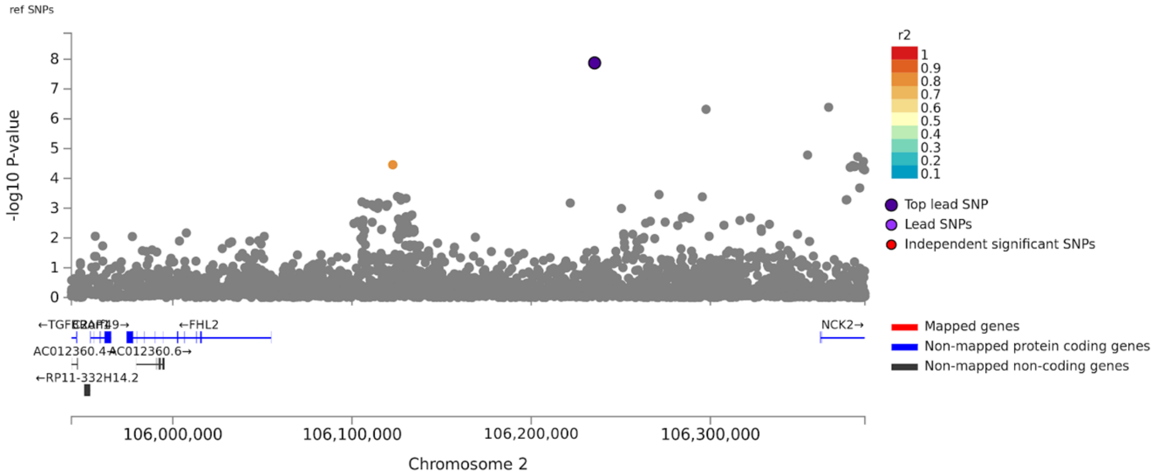


*Supplementary Figure 15*: Forest plot of rs115186657 highlighting the effect estimate (BETA), standard error (SE) and *P-value* of this variant in each dataset. The error bars represent 95% confidence intervals.

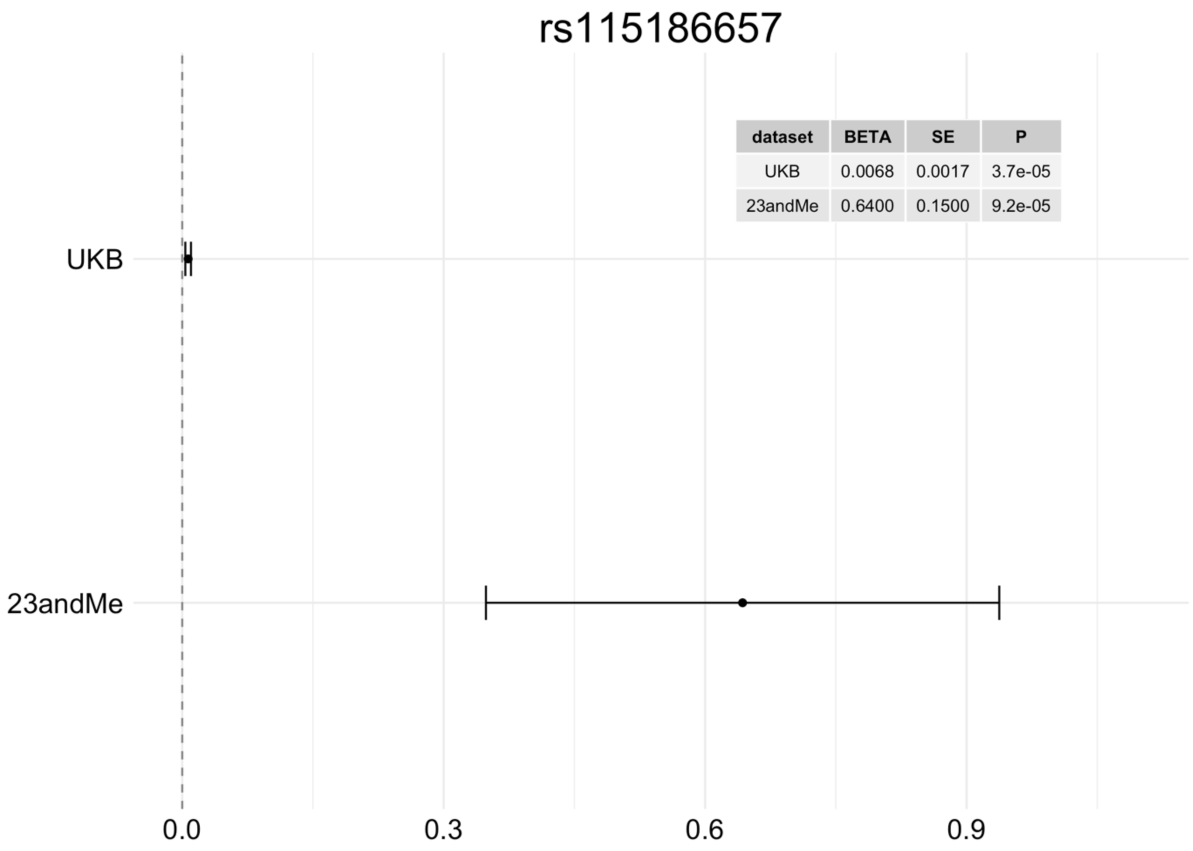


*Supplementary Figure 16*: Regional plot highlighting the lead variant of locus 7 (inconclusive) and the genes of interest. The eQTLs and the genes which they map to are included below the regional Manhattan plot.

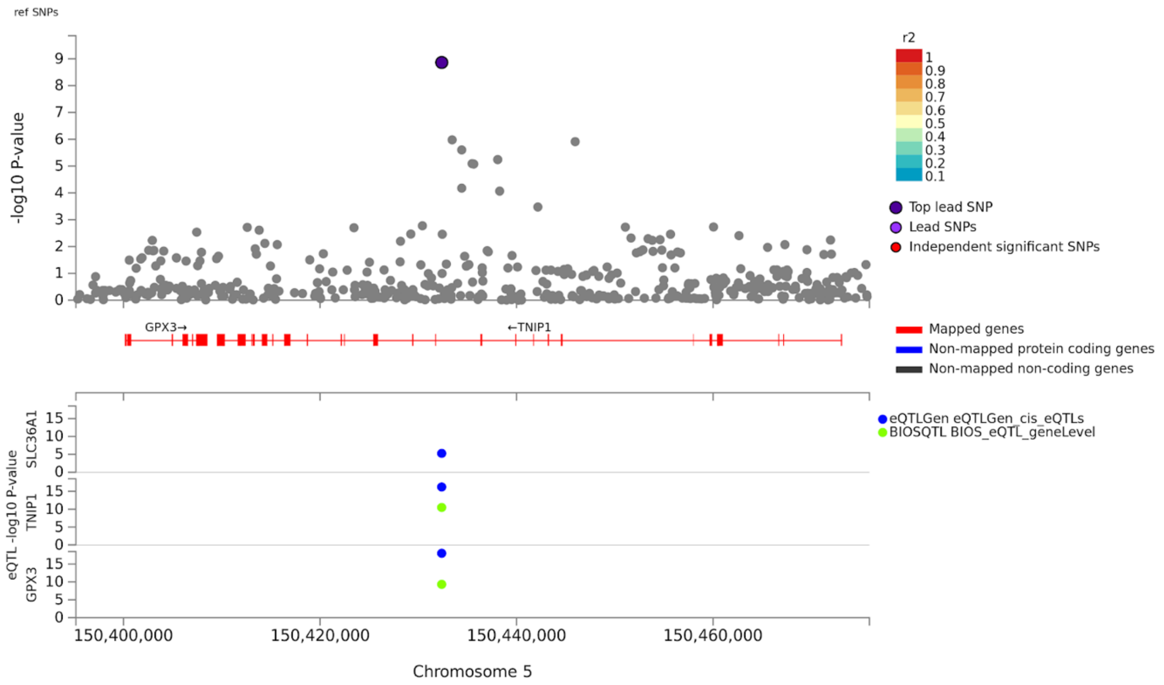

*Supplementary Figure 17*: Forest plot of rs871269 highlighting the effect estimate (BETA), standard error (SE) and *P-value* of this variant in each dataset. The error bars represent 95% confidence intervals.

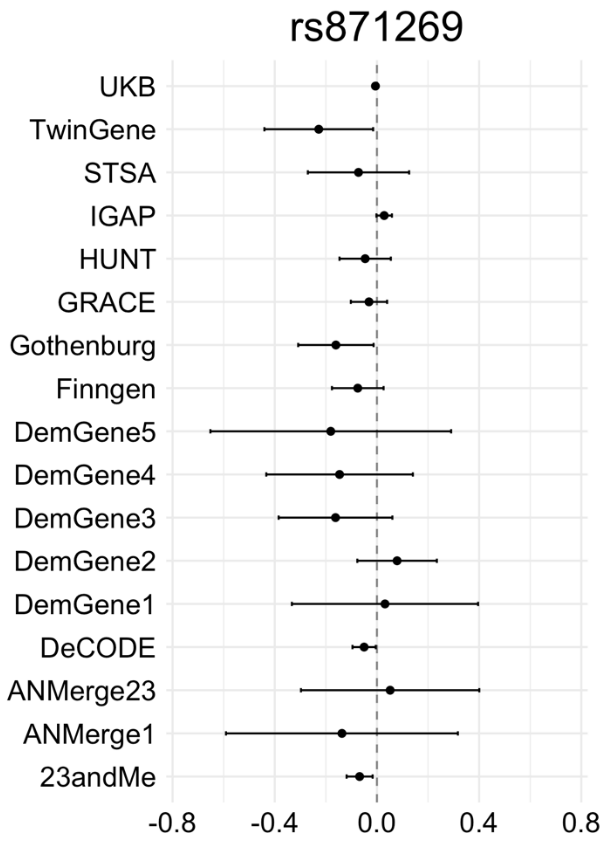


| dataset | BETA | SE | P |
|---|---|---|---|
| UKB | -0.0059 | 0.0016 | 0.00034 |
| TwinGene | -0.2300 | 0.1100 | 0.03600 |
| STSA | -0.0720 | 0.1000 | 0.48000 |
| IGAP | 0.0280 | 0.0160 | 0.06800 |
| HUNT | -0.0460 | 0.0510 | 0.37000 |
| Gothenburg | -0.1600 | 0.0750 | 0.03300 |
| GRACE | -0.0310 | 0.0360 | 0.39000 |
| Finngen | -0.0750 | 0.0520 | 0.15000 |
| DemGene5 | -0.1800 | 0.2400 | 0.45000 |
| DemGene4 | -0.1500 | 0.1500 | 0.32000 |
| DemGene3 | -0.1600 | 0.1100 | 0.15000 |
| DemGene2 | 0.0790 | 0.0790 | 0.32000 |
| DemGene1 | 0.0310 | 0.1900 | 0.86000 |
| DeCODE | -0.0500 | 0.0230 | 0.03100 |
| ANMerge23 | 0.0520 | 0.1800 | 0.77000 |
| ANMerge1 | -0.1400 | 0.2300 | 0.55000 |
| 23andMe | -0.0680 | 0.0260 | 0.00870 |

*Supplementary Figure 18*: The mean gene expression of 329 genes linked to risk loci through eQTL mapping across 64 brain regions highlights the top 10 regions with the highest expression. Gene expression values are obtained from the Allen Human Brain Atlas.

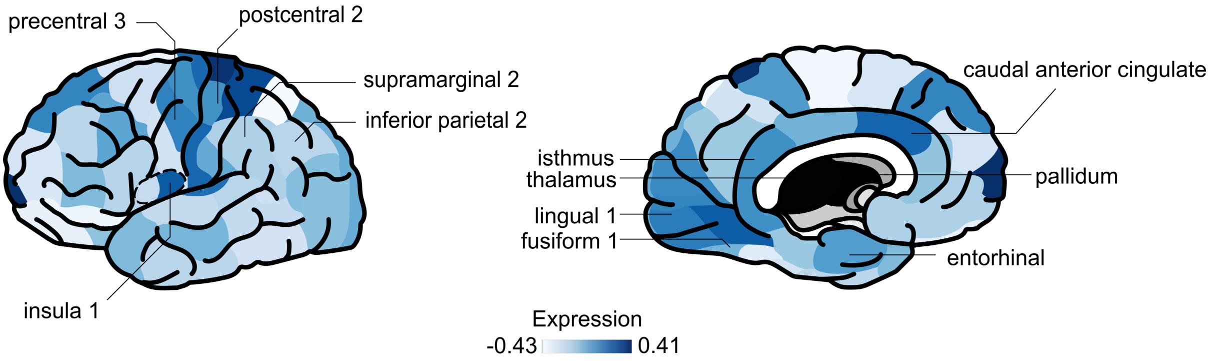

### Supplementary Methods

#### Datasets

##### deCODE

Data from the deCODE study included 7,002 Alzheimer's patients (5,098 of whom were chip-typed) and 181,573 controls (88,739 of whom were chip-typed). In 15% of patients, the diagnosis of Alzheimer's disease was established at the Memory Clinic of the University Hospital according to the criteria for definite, probable, or possible Alzheimer's disease of the National Institute of Neurological and Communicative Disorders and Stroke and the Alzheimer's Disease and Related Disorders Association (NINCDS-ADRDA). In 80% of patients, the diagnosis has been registered according to the criteria for code 331.0 in ICD-9, or for F00 and G30 in ICD-10 in health records. Five percent of the patients were identified in the Directorate of Health medication database as having been prescribed Donepezil (Aricept). The controls were drawn from various research projects at deCODE Genetics. The study was approved by the National Bioethics Committee and the Icelandic Data Protection Authority. Written informed consent was obtained from all participants or their guardians before blood samples were drawn. All sample identifiers were encrypted in accordance with the regulations of the Icelandic Data Protection Authority.

Chip-typing and long-range phasing of 155,250 individuals was carried out as described previously[35]. Imputation of the variants found in 28,075 whole-genome sequenced individuals into the chip-typed individuals and 285,664 close relatives was performed as detailed earlier[35]. Association analysis in the deCODE sample was carried out using logistic regression with AD status as the response and genotype counts and a set of nuisance variables, including sex, county of birth, and current age, as predictors[36]. Correction for inflation of test statistics due to relatedness and population stratification in this Icelandic cohort was performed using the intercept estimate (1.30) from LD score regression[37].

##### UK Biobank

The UK Biobank (UKB; www.ukbiobank.ac.uk)[38] summary statistics for 46,613 cases and 318,246 controls were obtained from Jansen *et al.* (2019)[3]. In short, a proxy phenotype for Alzheimer’s disease case-control status was generated from a self-report questionnaire which asked participants to report whether their biological mother or father ever suffered from Alzheimer’s disease/dementia, and to report each parent’s current age (or age at death, if applicable). The phenotype was constructed as a count of the number of affected parents ranging from 0 to 2. The contribution for each unaffected parent to the phenotype was weighted by the parent’s age/age at death. This was calculated as the ratio of parent’s age to age 100 (weight=(100-age)/100). The weight for an unaffected parent was capped at 0.32, corresponding to a risk equivalent to that of the maximum population prevalence of AD. Participants with a diagnosis of “Alzheimer’s disease” (code G30) or “Dementia in Alzheimer’s disease” (code F00) were given the maximum possible score of 2. Standard QC procedures were applied to the genotype data which was then imputed to the HRC[39], 1KG[2], and UK10K reference panels[40]. Further information on the quality control is available in Jansen *et al.* (2019)[3]. The imputed data was analyzed using linear regression with 12 ancestry principal components, age, sex, genotyping array, and assessment centre included as covariates. All participants provided written informed consent; the UKB received ethical approval from the National Research Ethics Service Committee North West-Haydock (reference 11/NW/0382), and all study procedures were in accordance with the World

Medical Association for medical research. Access to the UK Biobank data was obtained under application number 16406.

#### Nord-Trøndelag Health Study (HUNT)

The HUNT data consists of 1156 cases and 7157 controls, where cases were defined as individuals diagnosed with ICD-10 G30.0 or F00*, or ICD-9 331.0 and controls were individuals last seen as healthy with no previous diagnosis of Alzheimer's disease. All controls were more than 80 years old. Participants overlapping with the DemGene study were removed. Further information about the biobank is available at https://hunt-db.medisin.ntnu.no/hunt-db/#/.

The samples were genotyped with Illumina HumanCoreExome arrays (HumanCoreExome12 v1.0, HumanCoreExome12 v1.1, or UM HUNT Biobank v1.0). Participants with call rates <99%, contamination >2.5%, large chromosomal copy number variants, lower call rate of technical duplicate pair or twins, uncommon sex chromosomal conformations, or discrepancies in reported gender were removed. The samples passing QC were analysed in a second round of genotype calling, using the Genome Studio quality control described elsewhere[41]. Variants were mapped to the Genome Reference Consortium Human genome build 37 (http://genome.ucsc.edu) using BLAT[42].

Variants were excluded if they had call rates <99%, higher call rates in another assay, probe sequences not mapping to the reference genome, cluster separation <0.3, gentrain score <0.15, or Hardy Weinberg equilibrium deviation (*P-value*<0.0001) from unrelated European samples. We also removed variants with frequency differences > 15% between the datasets or that were monomorphic in one dataset and had MAF > 1% in one of the others. Only European ancestry individuals were included. Ancestry was inferred using PLINKv1.90[43], projecting the HUNT samples into the space of the principal components of the Human Genome Diversity Project panel[44,45]. The data was phased using Eagle2 v2.3[46], before imputing with Minimac3 v2.0[47], using a customized reference panel of HRC combined with 2,201 low-coverage whole-genome sequences HUNT samples. Variants with low estimated squared correlations between imputed and true genotypes ($R^2$ <0.3) were excluded. A logistic regression analysis was run with SAIGE[48], including sex, batch, and 4 PCs as covariates.

#### 23andMe

The 23andMe data consists of 3807 cases and 359,839 controls. Among the controls, there were 19,638 individuals between the age of 45-60 and 340,201 individuals over 60. There were 130 cases between 45-60 and 3677 cases over the age of 60. DNA extraction and genotyping were performed on saliva samples by National Genetics Institute (NGI), a CLIA licensed clinical laboratory and a subsidiary of Laboratory Corporation of America. Samples were genotyped on one of five genotyping platforms. The v1 and v2 platforms were variants of the Illumina HumanHap550+ BeadChip, including about 25,000 custom SNPs selected by 23andMe, with a total of about 560,000 SNPs. The v3 platform was based on the Illumina OmniExpress+ BeadChip, with custom content to improve the overlap with our v2 array, with a total of about 950,000 SNPs. The v4 platform was a fully customized array, including a lower redundancy subset of v2 and v3 SNPs with additional coverage of lower-frequency coding variation, and about 570,000 SNPs. The v5 platform, in current use, is an Illumina Infinium Global Screening Array (~640,000 SNPs) supplemented with ~50,000 SNPs of custom content. Samples that failed to reach 98.5% call rate were re-analyzed.

Only individuals of European ancestry were included in the data. Individuals were assigned ancestry by first partitioning the phased genomic data into short windows of about 300 SNPs. Within each window, a support vector machine (SVM) classified individual haplotypes into one of 31 reference populations (https://www.23andme.com/ancestry-composition-guide/). The SVM classifications are then fed into a hidden Markov model (HMM) that accounts for switch errors and incorrect assignments, and gives probabilities for each reference population in each window. Finally, we used simulated admixed individuals to recalibrate the HMM probabilities so that the reported assignments are consistent with the simulated admixture proportions. Europeans were defined as those with ancestry probabilities of European + Middle Eastern > 0.97 and European > 0.90. Only unrelated individuals were used for the GWAS analysis. Individuals were defined as related if they shared more than 700 cM IBD, including regions where the two individuals share either one or both genomic segments IBD. Cases were preferentially chosen over controls.

Variants were imputed in two separated imputation reference panels. For the first one, we combined the May 2015 release of the 1000 Genomes Phase 3 haplotypes[2] with the UK10K[40] imputation reference panel to create a single unified panel. We used the Human Reference Consortium (HRC) as the second imputation reference panel[39]. Participant data was phased using an internally-developed tool based on Beagle[49] and a new phasing algorithm Eagle[50]. The phased participant data was imputed against both reference panels using Minimac[40]. The resulting imputed data was merged with HRC given preference over the merged panel. The imputed dosage data was analyzed using age, sex, platform, and PCs 1-4 as covariates. The association test *P-value* was computed using a likelihood ratio test.

For QC of genotyped GWAS results, SNPs genotypes only on v1 and v2 platforms were flagged due to low sample size. SNPs on mitochondrial DNA and chromosome Y were flagged. Using trio data, SNPs that failed a test for parent-offspring transmission were flagged (specifically, child's allele count was regressed against the mean parental allele count and flagged SNPs with fitted $\beta<0.6$ and $P<10^{-20}$ for a test of $\beta<1$). SNPs with a Hardy-Weinberg $P<10^{-20}$, or a call rate of <90% were flagged. Genotyped SNPs with batch effects or date effects ($P<10^{-50}$) were flagged. SNPs with large sex effect (ANOVA of SNP genotypes, r2>0.1) were flagged. SNPs with probes matching multiple genomic positions in the reference genome ('self chain') were flagged. For imputed GWAS results, SNPs with Rsq < 0.3, as well as SNPs that had strong evidence of a platform batch effect were flagged. The batch effect test is an F test from an ANOVA of the SNP dosages against a factor representing v4 or v5 platform ($P<10^{-50}$). SNPs with a sample size <20% the total sample were flagged. These flagged SNPs were removed before analysis. Logistic regression results that did not converge due to complete separation, identified by abs(effect)>10 or stderr>10 on the log odds scale were removed. SNPs with MAF < 0.1% were removed.

#### BioVU

The BioVU data consists of 600 cases and 36,059 controls. Cases were defined as individuals diagnosed with ICD-10 G30 and ICD-9 331.0. Controls were individuals without any of the following ICD-10 diagnoses; G30, F01, F02, F03, F10.27, F10.97, F13.27, F13.97, F18.17, F18.27, F18.97, F19.17, F19.27, F19.97, G31.0, G31.83 and the following ICD-9 diagnoses; 331.0 ,290, 291.2, 292.82, 294.1, 294.10, 294.11, 294.2, 294.20, 294.21, 331.19, 331.82. Individuals with a family history of dementia in their electronic health records were also excluded from the control sample. The participants were genotyped on the Illumina MEGAEX array. The genotypes were filtered for SNP and individual call rates, sex discrepancies, and excessive heterozygosity using PLINK (--geno 0.05, --mind 0.02, |Fhet| > 0.2, HWE <$10x10^{-10}$). Autosomes were imputed to the HRC panel using Michigan Imputation Server1 in five batches and converted to hardcalls using default PLINK threshold settings. Non-biallelic SNPs were filtered out and SNPs with imputation quality (R2) less than 0.3. SNPs with minor allele frequency less than 0.005 were removed. SNPs with genotyping rates less than 0.98 were excluded. Individuals with call rates less than 0.98 were excluded.

Principal component analysis (PCA) was used to determine BioVU individuals of European genetic ancestry. First, we performed PCA using FlashPCA[51] on BioVU combined with CEU, YRI, and CHB reference sets from 1000 Genomes Project Phase 3[2]. Principal components were scaled so that the axes could be interpreted as proportions of genetic ancestry. We selected BioVU individuals who were within 40% of the CEU cluster along the CEU-CHB axis and within 30% of the CEU cluster on the CEU-YRI axis, generating a once-PCA filtered European set. To ensure subsequent steps would remove SNPs associated with reduced quality rather than cryptic population substructure, we filtered the previously identified BioVU European cluster to identify individuals falling within the CEU, TSI, and GIH 1000 genomes populations, producing a twice-filtered European set. Using the twice-filtered European set we conducted a series of SNP checks. We filtered individuals with IBS greater than 0.2. We checked for imputation batch effects by conducting pairwise logistic regression of the five imputation batches using sex and top 10 principal components as covariates. SNPs with p-values less than 0.001 in the additive model were flagged. Any SNPs with a MAF difference greater than 0.1 between BioVU and CEU were flagged. SNPs with a Hardy-Weinberg Equilibrium P-value less than $10x10^{-10}$ were flagged. Flagged SNPs were removed from the once-filtered European set. Individuals in the once-filtered European set and SNPs passing QC in the hardcall data were extracted from the dosage data. Finally, a GWAS was performed using SAIGE[48] with default settings including sex and the top 10 PCs as covariates.

#### DemGene, TwinGene, STSA, Gothenburg, and ANMmerge

The origin of the DemGene (1638 cases and 6059 controls), STSA (320 cases and 750 controls), and TwinGene (224 cases and 6321 controls) samples has been previously described in Jansen *et al.* (2019). For the STSA data, informed consent was obtained from all participants and the studies were approved by the Regional Ethics Board in Stockholm and the Institutional Review Board at the University of Southern California. For the TwinGene data, written informed consent was obtained from all participants and the study was approved by the Regional Ethics Board in Stockholm. The ANMmerge data (366 cases and 259 controls) consists of 3 batches. Batch 1 and 2 are available on synapse.org (synapse ID: syn22130010); the origin and genotyping of this data is described in Birkenbihl *et al.* (2020)[52]. Batch 2 and 3 were both genotyped on Illumina HumanOmniExpress-12 v1.0 and were merged after QC and removal of non-EUR individuals. The merged version of batch 2 and batch 3 were put through the same QC pipeline again and the batch associated variants were removed. Batch associated variants ($P<5x10^{-8}$) were identified through assigning the batch 2 individuals as controls and batch 3 individuals as cases and running Plink logistic regression.

The Gothenburg AD cases originate from Sweden and were either collected in memory clinics (in different parts of Sweden) or as a part of two population-based epidemiological studies in Gothenburg; the Prospective Population Study of Women (PPSW) and the Gothenburg Birth Cohort Studies (H70, H85 and 95+), described in detail previously [53–56]. Controls originate from the Gothenburg Birth Cohort Studies and PPSW. Individuals of non-European descent were excluded as part of the QC or the GWAS-data. AD diagnosis was based on National Institute of Neurological and Communicative Disorders and Stroke-

Alzheimer's Disease and Related Disorders (NINCDS-ADRA) criteria. All control samples were clinically investigated and free from dementia. The individuals were genotyped using the Illumina Neurochip array.

Initially, the genotype data were obtained in Plink v1.90[27] binary format and, if necessary, were converted to build GRCh37 using the UCSC LiftOver tool[28]. The raw genotypes were processed using the Psychiatric Genomics Consortium (PGC) Ricopili pipeline version 2019_Aug_16.001. The quality control (QC) procedure initially removed SNPs with a missingness > 0.95, then kept individuals with a SNP missingness < 0.05 and an autosomal heterozygosity deviation ($F_{het}$) < 0.2. Finally, SNPs with a missingness > 0.02, a difference in SNP missingness between cases and controls > 0.02; and deviation from Hardy-Weinberg equilibrium ($P < 10^{-6}$ in controls or $P < 10^{-10}$ in cases) were removed.

Next, non-European individuals within the datasets were removed based on principle component analysis (PCA), using the 1KG Phase 3 dataset as a reference[29]. The PCA pipeline was repeated including all European individuals in all genotype level datasets to identify individuals across the datasets with a pihat > 0.2 for exclusion from the analysis. PCA was additionally performed within each European dataset to create principal component covariates for logistic regression. The genotype data of the European individuals were imputed to the Haplotype Reference Consortium reference (HRC r1.1 2016)[30] using the Michigan Imputation Server[31]. The imputed genotypes within each dataset were analysed using Plink[27] logistic regression adjusted for covariates. The covariates were principal components 1-4, plus principal components significantly associated with the phenotype, and sex. The significant principal components were identified by testing the first 20 principal components for phenotype association and evaluating their impact on the genome-wide test statistics using λ. At the time of analysis age information was not available for use as a covariate however the cases and controls of each dataset were well matched in age (**Supplementary Table 15**). Unfortunately, only a fraction of age information for DemGene participants (40.4% of cases and 9.19% of controls) was available so age matching cannot be determined.

#### IGAP

The summary statistics from the International Genomics of Alzheimer's Project (IGAP)[57] were obtained from https://www.niagads.org/datasets/ng00075. The stage 1 results were used in the meta-analysis. The stage 1 results were derived from genotyped and imputed data (11,480,632 variants, phase 1 integrated release 3, March 2012) of 21,982 Alzheimer's disease cases and 41,944 cognitively normal controls. Further information on the methods for generating the summary statistics and phenotyping are available in Kunkle *et al.* (2019)[58]. The data was generated using standard QC procedures. The genotypes were imputed to the 1KG reference panel[2], analyzed with general linear mixed effects models and then meta-analyzed with METAL[59]. Written informed consent was obtained from study participants or, for those with substantial cognitive impairment, from a caregiver, legal guardian or other proxy, and the study protocols for all populations were reviewed and approved by the appropriate institutional review boards.

#### Finngen

The summary statistics for 1798 cases and 72206 controls from Finngen were obtained from https://storage.googleapis.com/finngen-public-data-r3/summary_stats/finngen_r3_AD_LO_EXMORE.gz. The genotype data were quality controlled with a standard protocol, imputed to SISu v3 reference panel, and analysed using SAIGE[48]. Thorough documentation of data sourcing and processing is available at https://finngen.gitbook.io/documentation/. Cases were defined as being diagnosed with ICD-10 G301, further information regarding the phenotype is available at https://risteys.finngen.fi/phenocode/AD_LO.

## GR@CE

The GR@CE data from Moreno-Grau *et al.* (2020)[60] was obtained through the GWAS catalog portal (ftp://ftp.ebi.ac.uk/pub/databases/gwas/summary_statistics/Moreno-GrauS_31473137_GCST009020/GRACE_StageI.txt). The phenotype was determined through structured neurological evaluation. The data was quality controlled using standard procedures, the genotypes were imputed to the HRC reference panel[39], and the dosages were analysed using an additive model in PLINK v1.9[43] with the top 4 PCs as covariates. Further information is available in Moreno-Grau *et al.* (2020)[60].

#### Brain regional gene expression

The per-region mean gene expression of the genes that map to the genomic risk loci based on eQTL expression was calculated using GAMBA (alpha version)[29]. A full description of the methods of GAMBA is available in Wei *et al.* (in preparation). In short, the gene expression data was obtained from the Allen Human Brain Atlas (http://human.brain-map.org) and the tissue samples were mapped to 64 FreeSurfer cortical and subcortical brain regions to generate a mean regional expression for each gene. Linear regression was used to compare the regional expression of the tested gene-set and the regional expression of the null model gene-set. The tested gene-set included 329 genes that mapped to genomic risk loci based on eQTLs and were present in the GAMBA gene expression data. The brain gene null model gene-set was composed of 329 randomly selected genes which are significantly over-expressed in the brain compared to other GTEx tissues. The random gene model was composed of 329 randomly selected genes with regional gene expression values in GAMBA.
